# Accurate and rapid turnaround of four hours for diagnosis of complicated UTIs using metagenomics

**DOI:** 10.1101/2025.04.24.25326310

**Authors:** Anurag Basavaraj Bellankimath, Sverre Branders, Isabell Kegel, Jawad Ali, Fatemeh Asadi, Truls E. Bjerklund Johansen, Can Imirzalioglu, Torsten Hain, Florian Wagenlehner, Rafi Ahmad

**Affiliations:** Department of Biotechnology, University of Inland Norway, Holsetgata 22, 2317, Hamar, Norway; Institute of Medical Microbiology, Medical Microbiome-Metagenome Unit (M3U), Justus Liebig University Giessen, Giessen, Germany; German Center for Infection Research (DZIF), Partner Site Giessen-Marburg-Langen, Giessen, Germany; Clinic for Urology, Pediatric Urology and Andrology, Justus Liebig University Giessen, Giessen, Germany; Institute of Clinical Medicine, University of Oslo, Norway; Institute of Clinical Medicine, University of Aarhus, Denmark; Institute of Clinical Medicine, Faculty of Health Sciences, UiT - The Arctic University of Norway, Hansine Hansens veg 18, 9019, Tromsø, Norway

## Abstract

Urinary tract infections (UTIs) affect 405 million people worldwide. Current diagnostics for UTIs rely on cultures, which can take 2 to 4 days. This study evaluated eleven culture independent methods for sample preparation from 77 complicated UTI patient samples, followed by real-time nanopore sequencing and data analysis. The metagenomic results were highly consistent with culture-based clinical routines. The optimized in-house method demonstrated a mean accuracy score of 99% for pathogen identification and 90% for antimicrobial susceptibility profiling. The method’s robustness is highlighted by accurately identifying the pathogen with as few as 33 bacterial cells/µL and a low bacterial-to-leukocyte ratio limit of 0.04.

Additionally, mNGS identified 13 pathogens that routine diagnostics missed, which were subsequently confirmed by Vivalytic or PCR. This method is up to 30% cheaper than published studies and commercial kits. AUROC analysis indicated that DNA yield and flow cytometry can be used for pre-screening to reduce costs, which is crucial for clinical adoption.

This research highlights the rapid diagnosis of UTIs in clinical settings using a cost-effective and scalable method that requires four hours from sample collection to informed decision making. Furthermore, it aims to improve antimicrobial and diagnostic stewardship by reducing empirical treatment and ensuring more judicious antibiotic use.

## Introduction

Urinary Tract Infections (UTIs) are the second most prevalent infections worldwide, affecting 405 million people annually and causing significant morbidity and economic costs. Over the past three decades, global deaths due to UTIs have increased dramatically by 140%^1,^ Untreated UTIs can progress to pyelonephritis^2^ and urosepsis, which accounts for a quarter of all sepsis cases^3, 4^. If not promptly treated, around 40% of hospital-acquired UTIs might cause these severe conditions^5^, posing a significant risk to immunocompromised, catheterized, and elderly patients. In severe UTIs, administering effective antibiotics as soon as possible is paramount.

However, a diverse range of virulence factors and pathogenic strains resistant to multiple drugs contribute to the complexity of UTIs. Rapid and accurate identification of pathogens and determination of antimicrobial susceptibility are central to managing UTIs and improving antimicrobial stewardship in an era of increasing antimicrobial resistance (AMR). Furthermore, AMR may be attributed to polygenic traits, which cannot be adequately identified through exclusive amplification of individual resistance genes. Given the advancing complexity of syndromic infections, polymicrobial infections, and AMR, the demand for multiplexed diagnostics has become increasingly important^6,7^. For culture-based methods, the time interval from sample collection to antibiotic susceptibility results is typically 2-4 days in the clinical routine^8, 9^.

Culture-free metagenomics next-generation sequencing (mNGS) can detect the complete metagenome (in both mono and polymicrobial UTIs) and known resistance mechanisms (AMR genes, SNPs, indels) present in a clinical sample. mNGS is also superior in identifying slow-growing pathogens and directly detecting AMR genes without a targeted assay. Studies in which the mNGS was applied directly to clinical urine showed concordance between the results of mNGS and conventional screening methods^10^. Processing to enrich target bacterial cells from patient specimens is crucial for developing effective sequencing-based diagnostics. Studies have determined the efficiencies of different extraction kits on patient samples such as stool and blood^11-13.^

However, efficient culture-independent methods for processing patient urine samples are rarely explored due to low bacteria-to-host cell ratios and the presence of crystal salts, urea, and β -hCG in urine, which complicates DNA extraction^14^. Furthermore, the human genome has approximately 3.05 billion bases^15^ compared to a typical bacterial genome, which has only 5 million on average^16^. Hence, the nucleic acid content of bacteria accounts for only a fraction (0.1%) of the human genome, thereby complicating the extraction of bacterial DNA from patient urine samples, which usually have high levels of host background ^17^. This requires implementing host depletion steps to enhance pathogen detection through metagenomics. Published studies are limited and have various drawbacks, such as focusing on uncomplicated UTI samples only, the need for large urine volumes, poor antibiotic susceptibility predictions, and longer turnaround times, resulting in higher costs per sample^18,19^. Additionally, mNGS can be performed for the same price as multiplex PCR, suggesting that the uptake of sequence - based methods is expected to increase.

This study aimed to develop a rapid and cost-effective method for accurately diagnosing complicated UTIs in clinical settings. Our objective was to evaluate 11 different techniques for culture-free DNA extraction directly from the urine samples of patients with complicated UTIs. This was followed by real-time nanopore sequencing and data analysis to identify pathogens and assess their antibiotic susceptibility. Such a culture- and amplification-independent workflow could transform the treatment of complicated UTIs.

## Results

Clinical samples present significant challenges for implementing metagenomic pipelines due to variable pathogen loads and the high ratio of the host to bacterial cells. Thus, selectively depleting the host enhances metagenomic methods. We developed eight in-house methodologies designed to selectively deplete abundant host cells and enrich bacterial populations (Figure 1 and Supplementary Table 1) and then compared the performance of these methods with three commercial test kits, starting with urine samples spiked with UTI-relevant bacteria followed by urine samples from patients with complicated UTIs. The optimized best performing method was further evaluated on additional patent samples. The mNGS results were compared with thos from conventional culture-based methods.

**Figure 1:**
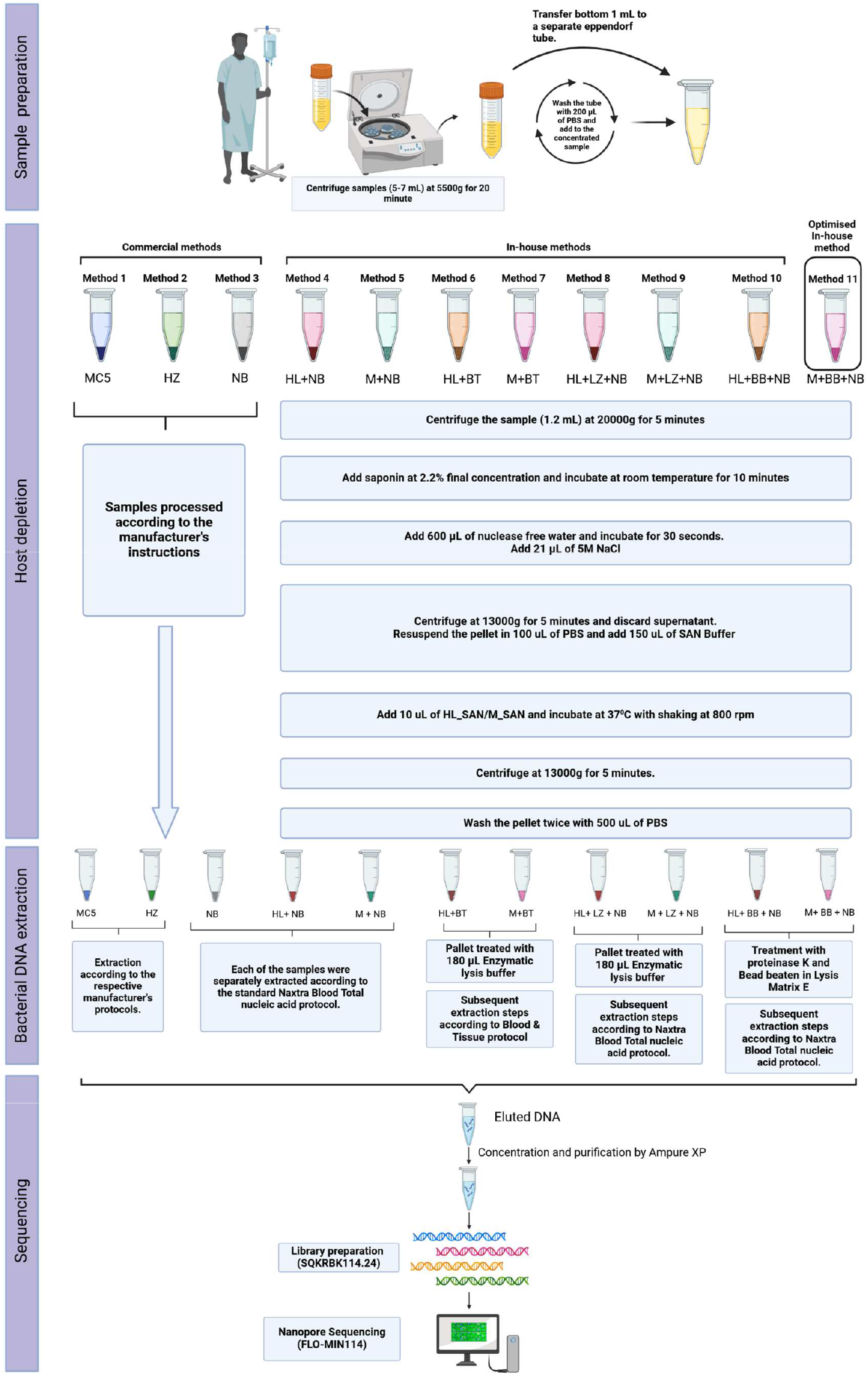
Schematic overview of various sample processing methods for clinical samples. Methods -3 are commercial, whereas methods 4-11 are SAN-based, in-house-developed techniques. Library preparation was performed using the rapid barcoding kit SQKRBK114.24, and the samples were sequenced on R 10.4.1 flow cells. The optimized method (M+BB+NB) performed best among the inhouse methods. Abbreviations: MC5: Molysis Complete 5, HZ: Host Zero Microbial DNA kit, NB: Naxtra Blood Total Nucleic Acid kit, HL+NB: HL_SAN + Naxtra Blood Total Nucleic Acid kit, M+NB: M_SAN + Naxtra Blood Total Nucleic Acid kit, HL+BT: HL_SAN + Blood and Tissue kit, M+BT: M_SAN + Blood and Tissue kit, HL+LZ+NB: HL_SAN + Enzymatic Lysis + Naxtra Blood Total Nucleic Acid kit, M+LZ+NB: M_SAN + Enzymatic Lysis + Naxtra Blood Total Nucleic Acid kit, HL+BB+NB: HL_SAN + Bead Beating + Naxtra Blood Total Nucleic Acid kit, M+BB+NB: M_SAN + Bead Beating + Naxtra Blood Total Nucleic Acid kit.

### Development of in-house methodologies

#### DNAyield-The in-house method showed similar performance to the commercial kits when tested on spiked urine samples

Eight in-house methods, which combined different heat-liable salt activated endonuclease (HL_SAN) and medium salt active endonuclease (M_SAN), were initially tested using urine spiked with *Escherichia coli*, along with four commercial methods as references (Supplementary Figure 1 and Supplementary Table 1). Among the in-house methods, DNA extracted with the Naxtra Blood (NB) in HL_SAN and M_SAN combinations, on average, yielded higher DNA (5.9 µg) than Blood & Tissue (BT) (3.5 µg). For the reference commercial methods, the NB kit (which doesn’t deplete host DNA) achieved the highest DNA yield at 10.7 µg, while the Host Zero (HZ) kit had the lowest at 1.0 µg. The results from the nanopore sequencing of the extracted DNA indicated very similar numbers of *E*.*coli* reads (>96.5%) across all the tested methods. These findings demonstrated that the in-house method performed comparably to the commercial kits regarding DNA yield and their compatibility and accuracy with nanopore sequencing (Supplementary Figure 1).

#### Host depletion - M_SAN shows slightly better performance in host depletion than HL_ SAN

For selecting among the salt-activated nuclease (SAN) enzymes, qPCR was performed to assess host depletion under different pH conditions using healthy urine samples spiked with *Enterococcus faecalis*. The results showed comparable depletion performance between M_SAN and HL_SAN, with M_SAN demonstrating a slightly higher depletion rate. Moreover, a slight loss of the spiked *E. faecalis* DNA was observed in the HL_SAN samples tested at pH 8.5 (Supplementary Figure 2).

**Figure 2:**
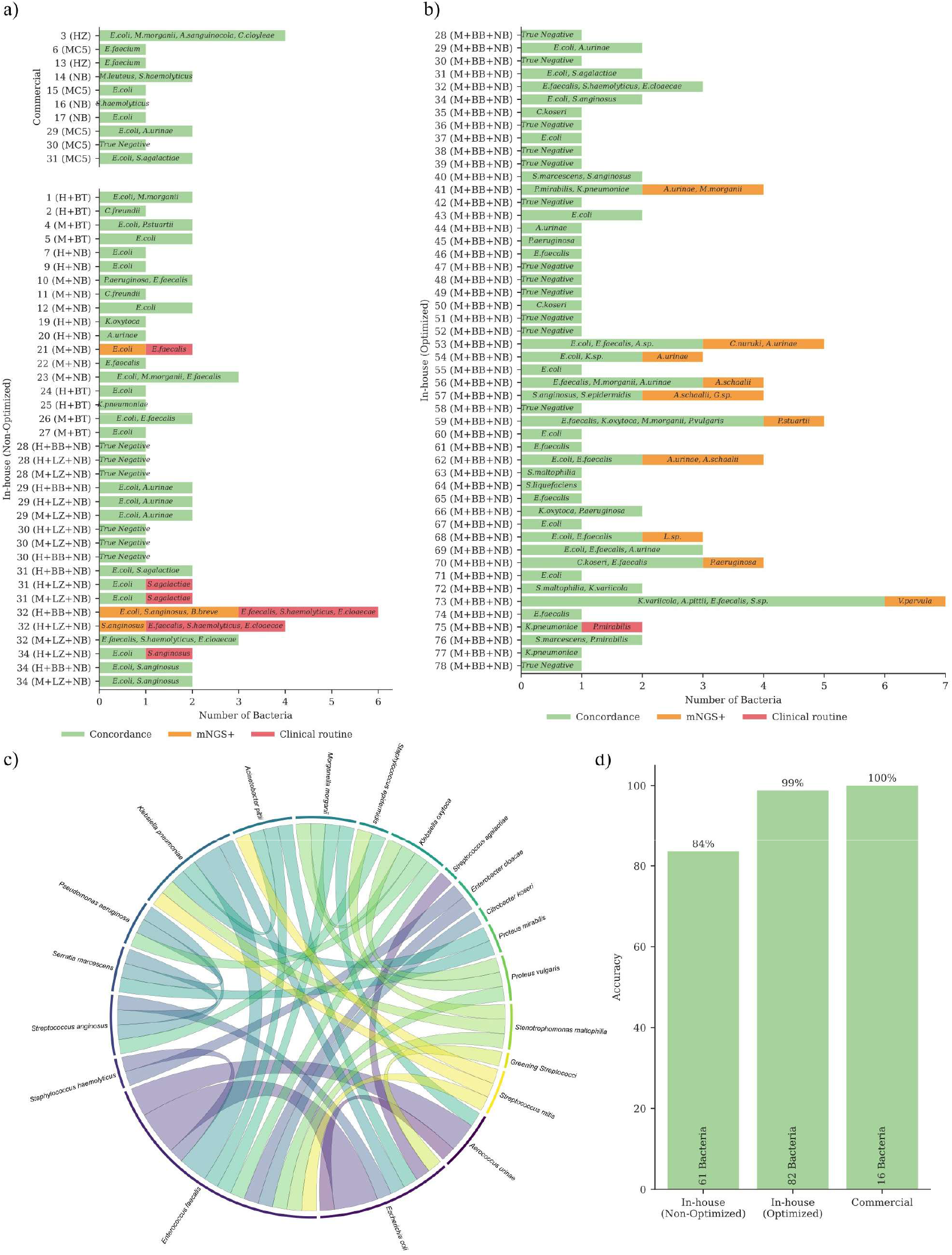
Overview of pathogen identification results. Subfigures (a) and (b) present bar graphs that indicate the species identified in each sample. Subfigure (a) displays samples extracted using Commercial and In-house (non-optimized) methods, while subfigure (b) illustrates the results for samples extracted using the In-house optimized method (M+BB+NB). Green bars represent species identified through routine testing and mNGS (concordant). Orange bars highlight those species uniquely identified by mNGS, and red bars denote species uniquely identified by routine testing. Subfigure (c) provides a chord diagram depicting the co-occurrence of pathogens in polymicrobial samples processed via the In-house optimized method. The ribbon’s thickness reflects how frequently the connected species appear in the same sample. Subfigure (d) features a bar graph of the overall accuracy (measured as prevalence threshold) of pathogen identification through mNGS across the three different methods, where routine identification is deemed the ground truth. The total number of bacteria considered for the accuracy calculation per method is annotated at the bottom of the bars.

#### The optimized in-house method demonstrates superior host depletion efficiencies compared to commercial methods

With M_SAN performing slightly better than HL_SAN, we evaluated the overall method performance in depleting the host to the two commercial kits, MC5 and HZ. NB was not tested as it lacks a host depletion step. qPCR assays were conducted to quantify depletion efficiencies, expressed as fold change. Using the optimized in-house method for the *E*.*coli* samples, we observed approximately 10^3^-fold depletion of human DNA, greater than HZ (approximately 10^2^-fold) and MC5 methods (approximately 10^1^-fold). No loss of bacterial DNA was observed in samples extracted using the optimized in-house method and HZ. However, a 2.2-fold loss of bacterial DNA (Average ΔCt=l .53) was found between the depleted and undepleted samples for MC5 (Supplementary Figure 3.a). Similarly, the optimized method outperformed others for samples spiked with *E*.*faecalis*, demonstrating a 10^2^-fold depletion of the host compared to HZ and MC5, which showed minimal to no significant depletion among those samples (Supplementary Figure 3.b). A slight loss of *E*.*faecalis* DNA was also noted in the samples extracted using HZ (Average ΔCt=l .55) and MC5 (Average ΔCt=2.93).

**Figure 3:**
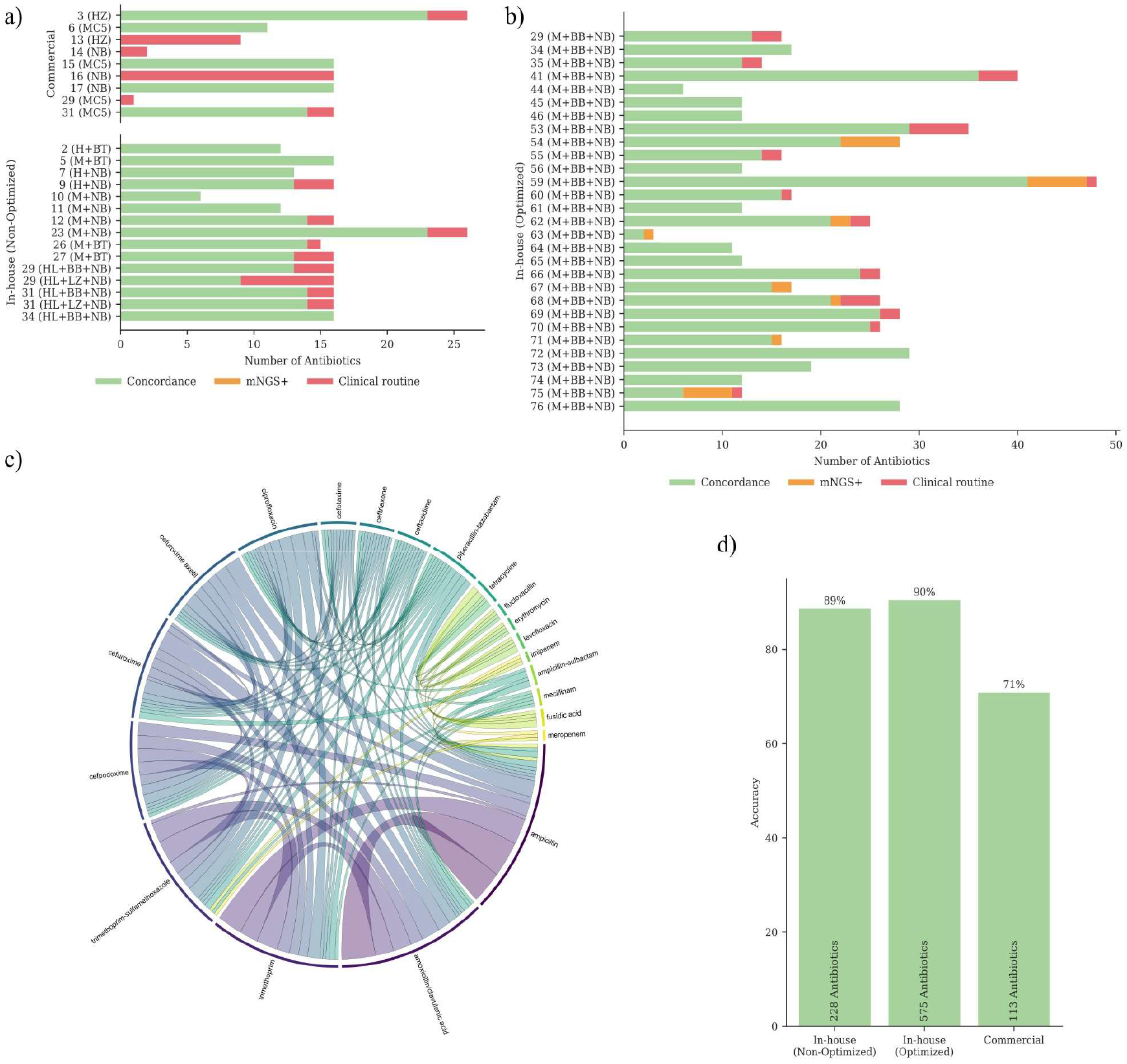
Overview of antimicrobial susceptibility results. Subfigures (a) and (b) present bar graphs that indicate the number of tested antibiotics per sample. Subfigure (a) displays samples extracted using Commercial and In-house (non-optimized) methods, while subfigure (b) illustrates the results for samples extracted using the In-house optimized method. Green bars represent concordance results between routine AST and mechanisms detected by mNGS for each antibiotic. Red indicates cases where resistance was identified in routine AST, but no corresponding resistance mechanism was detected through mNGS (false negative). Orange bars denote detected ARGs without phenotypic resistance to the corresponding antibiotic. Subfigure (c) presents a chord diagram illustrating the co-occurrence of antibiotic resistance in samples from the In-house optimized method, identified through routine testing. The ribbon thickness indicates how frequently the connected antibiotics exhibit resistance together in a sample. Subfigure (d) shows a bar graph depicting the overall accuracy (measured as the accuracy score) of antibiotic susceptibility testing through mNGS using the three different methods, with routine identification as the ground truth. The total number of species antibiotic combinations considered for accuracy calculation per method is annotated at the bottom of the bars.

## Comparison of mNGS vs. routine culture

### Routine microbiological findings underscore the complexity of UTI samples

All 77 samples collected for the method evaluation were leukocyte-positive based on the routine dipstick test. The routine microbiological culture results indicated that 82% (63) of the samples were culture positive, while 18% (14) were culture-negative. The positive samples were microbiologically complex, with 46% classified as polymicrobial and 54% as monomicrobial (Supplementary Table 2). Approximately 50% of the polymicrobial samples contained three or more pathogens, with five samples having as many as five (Supplementary Figure 4). Additionally, the samples exhibited complex resistance patterns, with 80% of tested samples resistant to at least one antibiotic. Of those resistant samples, 55% were multidrug-resistant. Therefore, a combination of both polymicrobial and multidrug-resistant UTI samples was evaluated across the methods.

**Figure 4:**
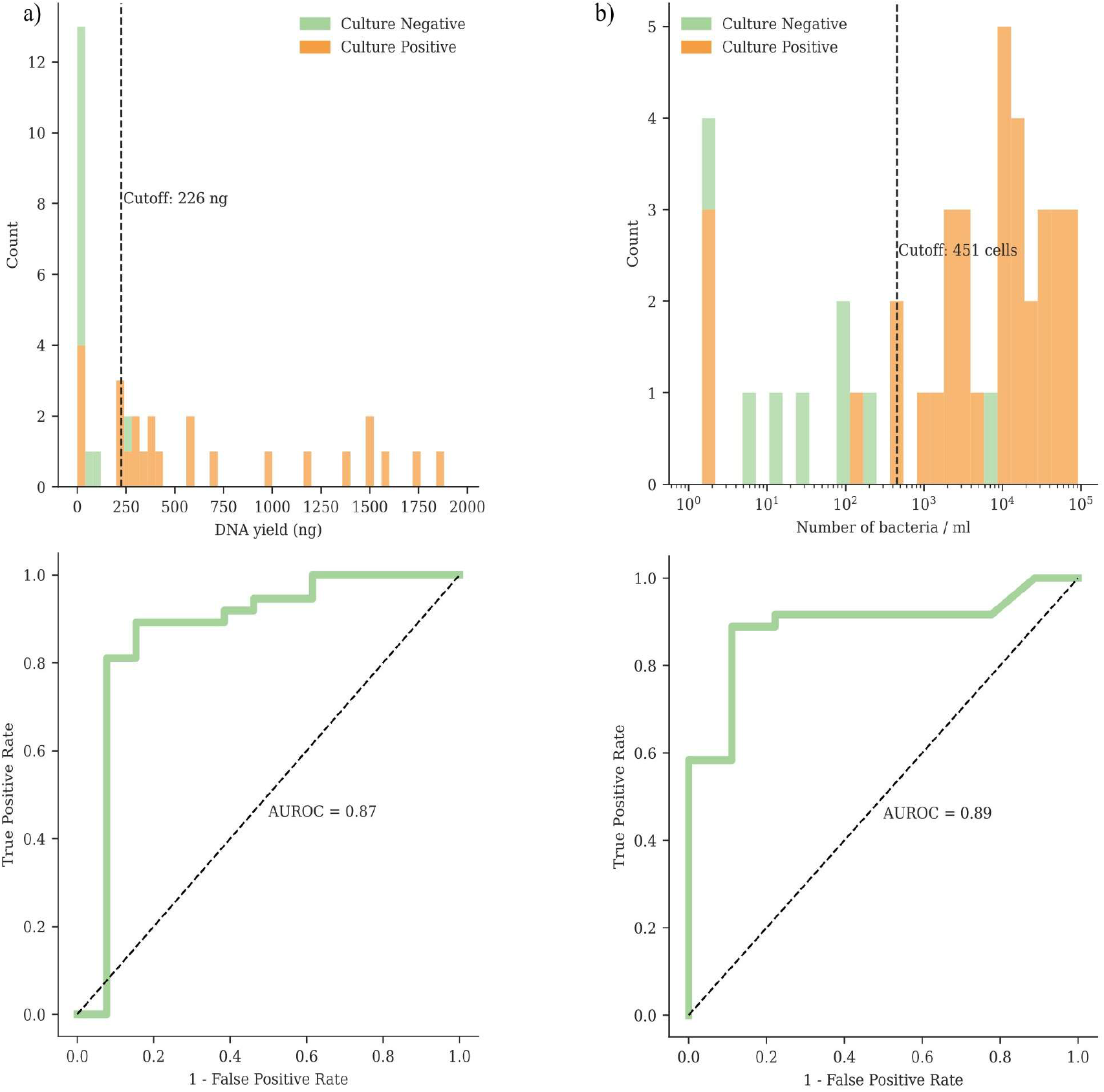
DNA yield and bacterial cell receiver operating characteristic curve analysis. The analysis was conducted on samples extracted using the in-house optimized method (n=50). Subfigure (a) presents the results for DNA yield, while (b) displays the results for the number of bacterial cells/mL as identified through flow cytometry. The histogram (top) illustrates the distribution of each variable for culture positive and negative samples, highlighting the optimal cutoff to distinguish between culture-positive and negative samples. The ROC curve (bottom) is annotated with the area under the ROC curve (AUROC) for each variable. The AUROC values of 0.87 (95% CI 0.70 - 0.98) and 0.89 (95% CI 0.78 - 0.99) demonstrate that both DNA yield and bacterial cell counts are reliable predictors of culture positivity.

#### Pathogen identification - the optimized method is more effective than other in-house methods and comparable to commercial methods

The metagenomic results closely matched the culture-based clinical routine in both pathogen identification and AMR detection. The three commercial methods (MC5, HZ, and NB) had 100% accuracy in identifying 16 pathogens across 10 samples. The non-optimized in-house methods demonstrated a mean accuracy score of 85% for pathogen identification across 61 pathogens and 33 tested samples (Figures 2a and 2d). Some of the non-optimized in-house methods could not detect some Gram-positive pathogens due to the inadequate performance of the NB method against these pathogens. In comparison, the optimized in-house method implemented a bead beating step and demonstrated a mean accuracy score of 99% across 75 pathogens and 50 samples (Figure 2b and 2d). The only false negative in the optimized method was sample 75, where *Proteus mirabilis* was not detected in a mixed infection with *Klebsiella pneumoniae. E*.*coli* and *E*.*faecalis* were the most common pathogens identified in 40% and 25% of the samples. Three of the 24 (13%) polymicrobial samples tested with the optimized method contained *E*.*coli* and *E*.*faecalis*. Two samples were found with *E*.*faecalis* - *K. pneumoniae* co-occurrence, and two samples had *E*.*faecalis -Aerococcus urinae* co-occurrence (Figure 2c).

Moreover, the optimized method identified 13 additional pathogens in nine tested samples that routine identification had initially failed to identify (Figure 2b). Routine testing identified three polymicrobial samples containing *A. urinae*, while mNGS identified *A. urinae* in four additional polymicrobial samples (41, 53, 54, and 62). mNGS identified *Actinotignum schaalii* in three polymicrobial samples (56, 57, and 62) where routine culturing did not. mNGS identified three additional pathogens that the traditional culture-based method missed: *M morganii* in sample 41, *Providencia stuartii* in sample 59, and *P aeruginosa* in sample 70. A re-analysis of these samples by Vivalytic later confirmed the mNGS findings. This highlights the higher accuracy and in-depth pathogen identification potential of the mNGS approach.

#### Susceptibility testing - the in-house optimized method outperforms other methods in predicting antibiotic susceptibility

For antibiotic susceptibility predictions, the non-optimized in-house methods achieved an accuracy of 89% across 228 antibiotics tested (Figures 3a and 3d). The optimized in-house methods achieved an accuracy of 90% across 575 antibiotics tested. The commercial methods had a 71% accuracy rate in predicting antibiotic susceptibility across 113 tested antibiotics (Figures 3b and 3d). Most false negative/positive susceptibility predictions were from the diaminopyrimidine, fluoroquinolones, and cephalosporins classes of antibiotics. In addition to rapidly detecting pathogens and ARGs, mNGS detected relevant markers like virulence genes (e.g., *kdpDE* in *E*.*coli*, was detected in samples 7, 9, 17, 23, 27, 37, 62, 67, and 68) and identified mutations associated with any known resistance mechanisms (e.g., *vanR-P* in *E*.*faecalis* was detected in sample 6), even for antibiotics not routinely tested. While the correlation of ARG data to phenotypic resistance remains challenging, these results highlight the ability to identify valuable information about resistance mechanisms and bacterial virulence, even in mixed samples.

AcrAB-TolC and EmrAB-TolC were the most frequently observed resistance mechanisms identified in 33% and 22% of the pathogens, respectively (Supplementary Figure 5). The most common co-occurrence of resistance mechanisms was EmrAB-TolC with MdtEF-TolC (19%) (Figure 3c), all of which were found in *E. coli*. The most commonly identified antibiotic resistance was against ampicillin (89%) (Supplementary Figure 6). The most prevalent co - occurring resistance was ampicillin and amoxicillin/clavulanic acid (39%). Ampicillin resistance often co-occurred with trimethoprim resistance (29%) in *E*.*coli, M morganii*, and *P aeruginosa*.

**Figure 5:**
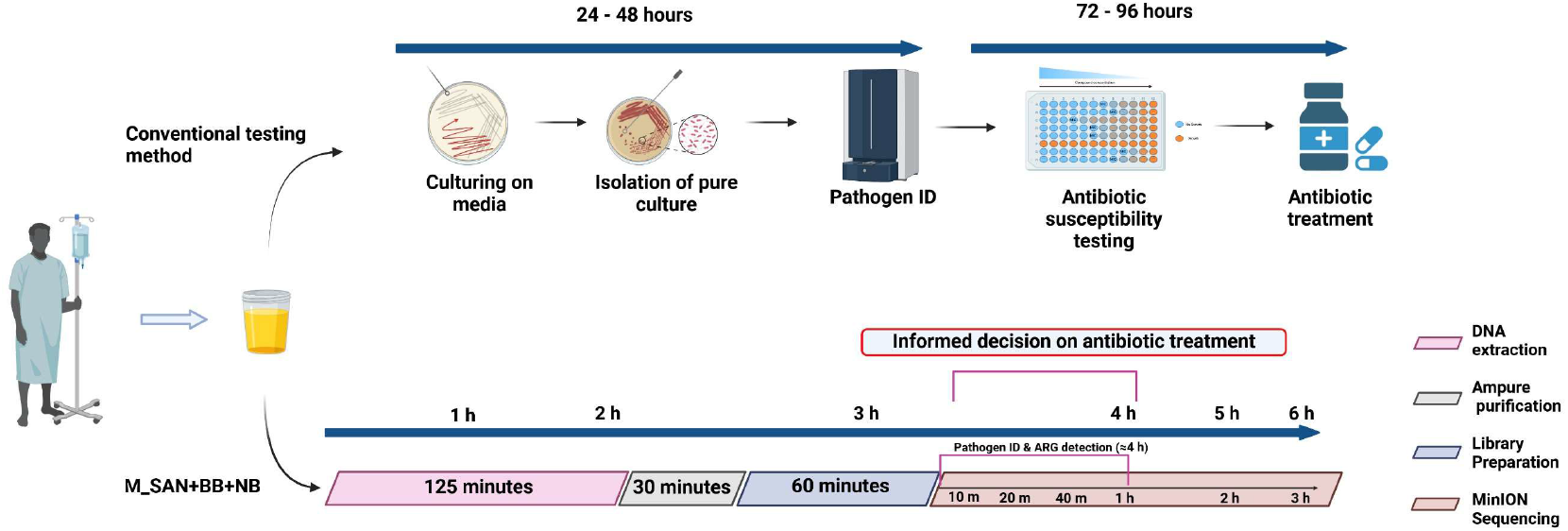
An overview of the timeline for pathogen and ARG detection using the in-house optimized method (M+BB+NB) compared to conventional routine diagnosis. The timeline for the method is organized and color-coded according to the specific steps involved: DNA extraction, purification, library preparation, and sequencing. This method has a four-hour turnaround time (TAT) from the sample receipt for detecting the pathogens and making informed decisions about antibiotic treatment.

#### Approximately 19% of unclassified reads were recovered using MysteryMaster

After sequencing, as much as 17% of sequencing reads longer than 500 bp remained unclassified after demultiplexing with Dorado. After processing the unclassified reads with the MysteryMaster demultiplex tool, it recovered, on average, 19% of those unclassified reads (Supplementary Figure 7). This recovery is particularly valuable for identifying pathogens in samples with either a low pathogen load, a very high leukocyte load, or a combination of both.

Bacterial identification also revealed that switching the basecalling mode from FAST to HAC or SUP resulted in an average increase of about 170% in identified bacterial reads. However, this increase did not influence the pathogen identification outcome, though it could enhance the information on AMR variants. Since no meaningful difference was observed in the number of bacterial reads between SUP and HAC modes, the SUP accuracy mode may not be warranted for bacterial identification. Therefore, using higher accuracy base-calling modes can only be recommended if abundant computational resources are available and time is not a limiting factor.

#### Strong relationship observed among flow cytometry data, DNAyield, and mNGS reads

Flow cytometry data about the number of bacterial and human cells (erythrocytes, leucocytes, round, and squamous epithelial cells) from clinical samples were used to estimate the host depletion by correlating cell numbers with the number of bacterial and human reads (Supplementary Figure 8). Sequencing data indicates that, on average, less than 1% of host reads were mitochondrial DNA, indicating depletion of both mitochondrial and host genomic DNA. Furthermore, the method’s robustness is demonstrated by its ability to accurately identify pathogens and predict AST in samples with a minimum bacterial-to-leukocyte ratio of 0.04 (Supplementary Figure 8).

An AUROC analysis was conducted to evaluate whether total DNA yield and/or the number of bacterial cells quantified through flow cytometry could serve as pre-screening indicators to differentiate between culture-positive and culture-negative samples (Figure 4). The analysis of DNA yield (Figure 4a) resulted in an AUROC of 0.87 (95% CI: 0.70 - 0.98). A cutoff value of 226 ng provided a positive predictive value (PPV) of 0.94 (95% CI: 0.88 - 1.00) and a negative predictive value (NPV) of0.73 (95% CI: 0.46-0.93). In comparison, the assessment of bacterial cell counts (Figure 4b) produced an AUROC of 0.89 (95% CI: 0.77 - 0.98), leading to a PPV of 0.97 (95% CI: 0.91 - 1.00) and an NPV of 0.67 (95% CI: 0.30 - 1.00) at a cutoff of 451 cells. The AUROC values indicate that both metrics are reasonable predictors of culture positivity. As this was a culture-independent study, the CFU/ml obtained through culturing was not a relevant criterion for AUROC calculations, as the CFU was obtained after 18-36 hours of incubation.

Moreover, the results imply a detection threshold of approximately 500 cells/mL, correlating with a positive predictive rate exceeding 90%, consistent with the clinical definition of a UTI. Further analysis indicated these cutoffs could be improved when stratified by sample origin (Supplementary Figure 9), i.e., whether the sample was a midstream urine sample or other (catheter, exprimate urine, first stream urine, etc.). DNAyield produced anAUROC of0.83 and 0.88 for midstream and other urine samples, respectively (Supplementary Figure 9a). The optimal cutoff values were 241 ng (PPV 0.96, NPV 0.71) and 18 ng (PPV 1.0, NPV 0.64). Bacterial cell counts (see Supplementary Figure 9b) produced anAUROC of 0.93 and 0.83 for midstream and other urine samples, respectively. The optimal cutoff values were 254 cells (PPV 0.96, NPV 1.0) and 39 cells (PPV 1.0, NPV 0.5).

### Time

#### Four hours of turn-around-time (TAT) from sample collection to pathogen identification and ARG detection

Testing the optimized method (M+BB+NB) on 50 clinical samples demonstrated that this method achieved the highest accuracy scores among all in-house developed methods, with 99% for pathogen identification and 90% for antimicrobial susceptibility predictions, making it the most effective extraction method. This method identified pathogens within 2 to 25 minutes of sequencing, while resistance genes were detected in as little as 3 minutes to a few hours. The turnaround time for the selected method, which includes host depletion, DNA extraction, library preparation, nanopore sequencing, and real - time analysis, was approximately 4 hours (Figure 5). In comparison, the commercial methods had turnaround times of 4.5 hours for MC5 and 6 hours for HZ (Supplementary Figure 10).

### Costs

#### The optimized method is also the most cost-effective

The method costs approximately $36 per sample, which includes DNA extraction, library preparation, and sequencing with a barcoding kit for 24 samples. These costs are lower than those of the commercial kits MC5 ($51) and HZ ($48). The sequencing cost was $31, covering the MinION flow cell and the rapid barcoding kit for 24 samples. This cost remained consistent across all methods, while the DNA extraction costs varied. The costs can be significantly reduced by pre-screening the samples based on the DNA yield and bacterial cell count cutoffs obtained from the AUROC analysis (Figure 4) since DNA extraction costs only about $6.

## Discussion

UTIs are among the most common bacterial infections, but diagnosing them can be challenging due to complex etiology and clinical presentations. Standard urine culture (SUC) has been the routine method for diagnosing UTIs for decades; its lengthy turnaround time and overall sensitivity are major limitations^20,21^. Unlike the SUC, metagenomics is a target-agnostic approach that facilitates rapid and targeted antibiotic treatment of UTIs^22^. In this study, we aimed to develop a rapid, selective, and cost-effective host depletion-based metagenomic workflow for analyzing UTI samples. Development comprised comparing the performance of these methods with commercial alternatives.

In-house methodologies were developed that combine saponin and SAN endonucleases for host depletion, along with magnetic bead- based (NB) or column-based DNA extraction (BT) methods. In the initial qPCR-based studies on spiked urine samples, M_SAN showed a slight advantage over HL_SAN in host depletion, likely because M_SAN functions best in a neutral to slightly basic pH environment, whereas HL_SAN requires a fully basic pH for optimal performance^23,24^. The magnetic bead-based NB method was preferred for DNA extraction due to several advantages over the BT method. Advantages include longer average read lengths (1,957 bp compared to 1,293 bp), shorter assay times (45 minutes instead of 78 minutes), lower per-sample extraction costs ($3.30 vs. $4.10), and potential for more straightforward automation. However, the NB method faced challenges with Gram-positive bacteria, prompting improvements in the lysis steps by incorporating enzymatic or bead-beating procedures in each SAN+NB variant. Furthermore, this optimized protocol (M+BB+NB) outperformed the commercial kits MC5 and HZ in depleting host DNA and enriching *E. coli* DNA in the spiked urine samples. A comparable outcome was observed with samples spiked with *E. faecalis*. Consequently, these spike-in experiments demonstrate that the in-house method effectively eliminated host DNA while preserving bacterial DNA.

Eleven methods, consisting of eight in-house methods and three commercial kits, were evaluated using 77 complex UTI patient samples. The urine samples were everyday clinical samples retrieved as midstream-, and catheter urines from patients in a reference hospital for complicated, severe UTIs. To our knowledge, no other study has evaluated such a wide variety of urine samples. The pathogen identification accuracy for the optimized in-house methods was 99%, while the commercial methods achieved 100%. Noteworthy, most reads (96 - 99%) from samples 14 and 16, extracted via the NB method, were host reads, underscoring the absence of a host depletion step in this method (see Supplementary Table 2). The overall accuracy of antibiotic susceptibility predictions varied among methods, with the optimized in-house method achieving an accuracy of 90% compared to 71% for the commercial methods. Predictions related to cephalosporins, quinolones, and diaminopyrimidines were the primary contributors to false predictions (Supplementary Figure 5). This was mainly linked to the detection of *ampC* variants and the efflux pumps AcrAB-TolC and MdtEF-TolC. Most of the detected *ampC* variants *(blaEC-5, blaEC-8, blaEC-15, blaEC-18, and blaEC-19)* were found in samples of *E. coli*. Although *E. coli* naturally possesses the chromosomal *ampC* gene, the absence of the *ampR* gene leads to reduced expression^25^. The low levels of expression of the *ampC* gene in *E. coli* may result from mutations in the promoter and attenuator regions rather than induction due to exposure to antibiotics such as β-lactams^26^. Hence, their detection rarely leads to relevant detectable resistance. Similarly, detecting efflux pumps AcrAB-TolC and MdtEF-TolC alone rarely results in significant phenotypic resistance^27^. In addition to detecting ARGs, mNGS identified relevant virulence genes such as *kdpDE* in *E. coli*^*28*^. The method identified mutations linked to known resistance mechanisms like *vanR-P* in *E. faecalis*, even for antibiotics not routinely tested. Despite the challenges in correlating ARG data to phenotypic resistance, these findings highlight the potential to uncover important information regarding resistance mechanisms and bacterial virulence, even in polymicrobial samples.

The optimized in-house method (M+BB+NB) detected all but one pathogen among the 50 tested clinical samples, identifying a total of 82 pathogens. This optimized method accurately identified pathogens in highly complex samples containing up to five unique pathogen species and a significant number of host immune cells. Furthermore, it identified additional pathogens in polymicrobial samples, aligning with findings from previously published work^30^. Overall, the optimized method had a pathogen identification accuracy of 99% and an antibiotic susceptibility prediction accuracy of 90%. Therefore, this approach achieves an overall accuracy rate exceeding 90%, which is typically essential for implementing new diagnostic techniques in clinical microbiology^31,32^. The method’s accuracy also compares well with existing studies on clinical samples. Pathogen identification concordance between the optimized method and clinical routine results (n=82, 99%, precision 83%, recall 99%) was higher than results reported in comparable studies: Liu et al., 2023 n=l 327 95% (precision 64%, recall 88%)^18^, Zhang et al., 2022 n=75 92% (precision 98%, recall 87%)^19^, Janes et al., 2022 n=81 89% (precision 87%, recall 79%)^30^, Jia et al., 2023 n=43 77% (precision 34%, recall 50%)^33^.

Almost half of the 50 sample cohorts used to evaluate the optimized method involved polymicrobial infections. Further analysis of these samples revealed a significant co-occurrence of *E. coli* and *E. faecalis*. This observation aligns with prior research, demonstrating that these two species frequently co-exist in polymicrobial UTI cases, especially in catheterized patients, where they co-localize within a biofilm on catheters and employ dynamic adaptive strategies that promote their coexistence^34,35^. Interestingly, *A. urinae* and *A. schaalii* were detected five and three times, respectively, in polymicrobial samples through mNGS, while routine culturing did not identify them^36,37^. Both pathogens are often underreported because they are difficult to identify, especially in polymicrobial samples, and are frequently dismissed as contaminants^38^. Furthermore, the method identified three additional pathogens, which the traditional culture based method missed. All of these samples were re-evaluated using the Vivalytic analyzer^39^, and the presence of the additional pathogens was confirmed. This highlights the power of the in-house method in detecting difficult-to-culture pathogens.

Moreover, published works typically evaluate AST concordance by antibiotic class (e.g., β-lactamases)^30^, if at all1^8,40^, rather than by individual antibiotic-pathogen combinations as in the current study. The optimized method achieved 90% AST concordance for 575 antibiotic pathogen combinations, including highly complex polymicrobial samples. Our work emphasizes the potential of the M+BB+NB method for AST prediction and pathogen identification in clinical UTIs across a wide range of sample complexities, with a limit of detection of 10^3^ CFU/ml, aligning with the clinical definition of UTI.

Among the samples analyzed, 80% of the positive ones were resistant to at least one antibiotic, and 55% exhibited resistance to multiple antibiotics. Additionally, 89% of resistant isolates showed resistance against ampicillin. This resistance frequently co-occurs with resistance to amoxicillin/clavulanic acid or trimethoprim. It has been suggested that co-selection via ampicillin and amoxicillin is a key factor driving trimethoprim resistance^41^. The most commonly identified resistance mechanisms among resistant samples were AcrAB-TolC and EmrAB-TolC. AcrAB-TolC was frequently found to co-occur with MdtEF-TolC. AcrAB is constitutively expressed in *Enterobacteriaceae*. EmrAB and MdtEF are differentially expressed under stress conditions, such as acidic environments, which may exert selective pressure on all three proton-motive force-driven efflux pumps ^42 - 44^.

The current clinical routine diagnostics ofUTI take 2-4 days (Figure 5), necessitating a lengthy regimen of empirical antibiotic treatment in severe cases. Decreasing this time could greatly promote prudent antibiotic use. While a mNGS based detection method with a TAT of as little as 5 hours has been proposed ^1^4,it was tested on spiked urine samples rather than clinical ones. Previously, Illumina-based mNGS methods have been tested on clinical UTI samples, but the sequencing time of these methods takes at least one day^30,33,45^. Nanopore-based methods are more rapid, and such methods with TATs ranging between 10 and 6 hours have been successfully tested on clinical UTI samples^18,19^. The current study further improves upon this, with the optimized method having a total TAT of only 4 hours (Figure 5). Timely treatment with the appropriate antibiotic is essential for effectively managing severe infections, such as urosepsis and surgical cases, and antibiotic stewardship.

Cost-effectiveness is of vast importance when adopting a technology that improves the treatment of patients and antimicrobial stewardship. The optimized method is cost-effective, costing about $36, which is 30% cheaper than the tested commercial kits. This cost includes DNA extraction, library preparation, and sequencing expenses for 24 multiplexed samples. When handling larger sample sizes, sequencing costs can be further reduced by multiplexing up to 96 samples and sequencing on a PromethION flow cell, lowering the cost to $15^46,47^. The overall per-sample costs can be further reduced by utilizing flow cytometry-based cell counts and DNA yield cutoffs. Previous studies have demonstrated that flow cytometry can detect significant bacteriuria in patient urine samples but lacks the capability to identify the pathogen^39, 48^. In this study, we built upon this observation and assessed the feasibility of using bacterial cell counts from flow cytometry data and DNA yield as pre-screening criteria to differentiate between culture-positive- and negative samples. The AUROC analysis showed 0.89 (95% CI 0.78 - 0.99) for bacterial cell count and 0.87 (95% CI 0.70 - 0.98) for DNA yield, indicating that both flow cytometry and total DNA yield can serve as indicators for distinguishing between positive and negative samples. This approach could significantly reduce costs to around $6, eliminating the expense of sequencing negative samples.

Furthermore, the sequencing results indicate low levels (<1% of host reads) of mitochondrial DNA, suggesting that the mitochondrial membrane is also lysed, resulting in DNA depletion. Previously published methods^21^ have struggled with selectively depleting mitochondrial membranes, leading to a significant amount (88.8%) of sequencing data of mitochondrial origin. Our optimized strategy has also successfully addressed this limitation.

A limitation of the developed host depletion protocols is their inability to detect fungal pathogens. The lysis effect of saponin is mediated through interaction with sterol groups present in eukaryotic cell membranes^49^ but not in bacterial cell membranes^50^. Unlike bacteria, fungal cell membranes contain sterol ergosterol, which increases their susceptibility to saponin and subsequent osmotic stress^51^. Therefore, a revised protocol should facilitate the detection of fungal pathogens. The optimized method (M+BB+NB) was tested on a limited number of samples in the present study. It is currently being evaluated with an additional 200 clinical urine samples to assess its robustness and improve the analytical metrics.

In summary, this study demonstrates the effectiveness of our optimized DNA extraction method (M+BB+NB), enhanced by nanopore sequencing and real-time data analysis, for the rapid diagnosis ofUTIs in clinical settings. Furthermore, these mNGS techniques can provide deeper insights into resistance mechanisms. An accurate, scalable, easy-to-implement, and cost-effective rapid point-of-care method can improve the clinical management of time-sensitive conditions, including urosepsis and kidney transplant infections. Additionally, it will strengthen antimicrobial and diagnostic stewardship by shortening the duration of empirical treatment and ensuring more judicious antibiotic use. By reducing the diagnostic timeframe from four days to four hours, this approach could prevent the unnecessary use of 1.62 billion daily doses (405 million cases × 4 days) of empirical broad-spectrum antibiotics each year before initiating personalized, targeted therapy against an identified pathogen. This will not only optimize UTI treatment but also enhance antimicrobial stewardship and mitigate further development of antimicrobial stewardship and mitigate further development of antimicrobial resistance (AMR).

## Methods

### Study design

We developed eight methodologies for complex UTI diagnostics using real-time nanopore sequencing and data analysis (Supplementary Table 1). Eleven methods, including eight combinations of SAN endonucleases (HL_SAN/M_SAN) and three commercial kits (Figure 1), were evaluated, with a total of 77 patient samples collected from the urology department. The study was conducted in three phases: a method development phase involving spiked samples and the clinical evaluation of the developed methods in two testing phases at a urology department. In the pilot testing phase, 11 different methods were initially tested on 34 patient samples (Figure 1). Afterward, the best-performing method was chosen based on a combination of host depletion, cost, accuracy, and turnaround time. The optimized method was tested on 50 additional samples to assess its performance compared to the microbiology routine’s clinical results.

### In-house method development with spiked urine

A saponin and SAN endonucleases (HL_SAN/M_SAN)-based assay was developed and tested on spiked urine samples. Saponin and SAN have been previously described for respiratory samples, including sputum, bronchoalveolar lavage, and endotracheal secretions^52^. The 3-10 ml urine sample was centrifuged at 5500g for 20 minutes at room temperature, after which the supernatant was discarded, leaving behind 1 mL at the bottom. After resuspending the bottom 1 mL and any possible pellet in an Eppendorf tube, the spent tube was rinsed with 200 µL of PBS. The resulting 1. 2 mL sample was centrifuged at 20000g for 5 minutes, and the top 500 µL of clear supernatant was discarded. The remaining 700 µL was used for subsequent steps. Then, 700 µL of 4.4% saponin solution (Sigma Life Sciences) was added to the sample, and the mixture was incubated in an Eppendorf ThermoMixer block for 10 minutes at 800 rpm. Next, 600 µL of nuclease-free water (NFW) was added to the mixture, vortexed, and incubated at room temperature for 30 seconds, followed by the addition of 21 µL of 5M NaCl to induce osmotic shock and lyse host cells. The mixture was then centrifuged at 13000g for 5 minutes, after which the supernatant was discarded, and the resultant pellet was resuspended in 150 µL of PBS. Subsequently, 150 µL of SAN buffer (5.5 M NaCl, 0.1 M MgCh) was added to the resuspended pellet with 10 µL of either HL_SAN (25K U/µL, Arctic-Zymes Technologies, Troms0, Norway) or M_SAN (292 U/µL, Arctic-Zymes Technologies, Troms0, Norway). The mixture was incubated at 37 ° C for 15 minutes at 800 rpm in an EppendorfThermoMixer. The samples were centrifuged at 13000g for 5 minutes, after which the supernatant was discarded, and the pellet was washed twice with 500 µL of PBS. The obtained pellet was then used for the extraction of microbial DNA. Following host depletion, the microbial DNA was extracted using the Naxtra Blood Total nucleic acid kits (LSBL 0048, Lybe Scientific AS, Norway, here referred to as NB) or the DNasey Blood and Tissue kit (cat: 69504, Qiagen, Germany, referred to as BT). The extractions were carried out according to the respective manufacturer’s instructions for both kits, and the Gram-positive pre-processing steps for BT were followed during the extractions.

### Commercial kits used as reference

In addition to the developed SAN assays, four different commercial DNA extraction kits,Molysis Complete 5 (D-321-050, Molzym GmbH, Germany, from here on referred to as MC5), Host Zero microbial enrichment kit (D4310, Zymo Research, USA here on referred as HZ), Naxtra Total nucleic acid (LSNXC0048, Lybe Scientific AS, Norway here on referred to as NT) and NB, were evaluated as standalone kits. The experiments were conducted according to the standard kit protocol without modifications.

### DNA yield and nanopore sequencing testing of the methods with spiked urine

For initial method development, healthy urine samples were spiked with *Escherichia coli* NCTC 13441 at 10^8^ and 10^5^ CFU/mL concentrations according to the previously established spiking protocols^53^. Dipsticks were used to confirm the healthy urine samples. Initially, the spiking was carried out in 5 mL of urine, but the volume was later increased to 10 mL. After spiking, the initial SAN assays included a 10% EDTA pre-treatment step to prevent crystallization and aid in DNA extraction. The extractions were performed with all eight SAN combinations (four SAN combinations extracted with NB and four SAN combinations extracted with NB pre-treated with 10% EDTA) and four commercial kits. Spiked pathogenic *E. coli* DNA was verified using PCR, as described previously^53^. A subset of the extracted DNA samples (commercial kits and HL_SAN/M_SAN methods extracted with NB) were sequenced on MiniON RI 0.4.1 flow cells.

### Comparison of the depletion efficiencies between HL_SAN and M_SAN endonucleases

Urine obtained from healthy donors was confirmed through a negative dipstick test and then spiked with WBC from buffy coat to mimic clinical samples. It was subsequently spiked with *E*.*faecalis* to achieve the final concentration of 10^5^ CFU/mL^53^. The WBC and bacterial spiking were performed on the main batch of healthy urine to establish a common baseline across all the aliquots. Following spiking, DNA was extracted using H+BT and M+BT methods for test samples, whereas the undepleted control samples were extracted using the standard Blood and Tissue protocol. All extractions were performed in triplicate (n=3), and comparisons were carried out using a qPCR assay to measure the effects on host DNA and E. faecalis DNA. The results were further analyzed to determine p-values among different replicate groups using a two-tailed unpaired t-test.

### Comparison of the depletion efficiencies of the optimized method to commercial kits

To determine their relative host depletion abilities, the in-house method M+BB+NB was compared to two commercial kits, MC5 and HZ. Similar to the endonuclease experiments, healthy urine samples were spiked with WBCs isolated from the blood and a uropathogen at 10^5^ CFU/mL to mimic clinical samples. The experiment was conducted in two batches: one spiked with *E. coli NCTC* 13441, and the other spiked with *E. faecalis* CCUG 9997. Spiking was carried out on the pooled batch in each instance to maintain a uniform baseline. The spiked samples were then tested in triplicate using the three methods to assess host depletion and any unintended loss of bacteria. SYBRgreen-based qPCR assays were performed to quantify the number of bacteria and hosts among the depleted and undepleted samples.

### qPCR assays

SYBRGreen-based qPCR assays targeting host DNA (human -actin gene) and *E. coli* (*UspA* gene) or *E. faecalis* (*GroES* gene) were performed on samples. Each of the qPCR assays included 3 µL of 5X HOT FIREPol® EvaGreen® qPCR Supermix (Solis BioDyne, Estonia), 0.2 µM of forward and reverse primers, 10.4 µL of nuclease-free water, and 1 µL of DNA template. The assays were carried out using a 7500 Fast Real-Time PCR system (Invitrogen ™, USA) using the following thermal profile: initial denaturation at 95°C for 720s followed by 40 cycles of amplification at 95°C for 25s, 60°C for 45s, 72°C for 60s. The extent of host depletion and bacterial enrichment was calculated using Ct, normalized using the Ct of the undepleted biological control. The Normalized Ct values were represented as fold changes calculated using equation 2-L’lCt_ The results were further analyzed to determine p-values among different replicate groups using a 2-tailed unpaired t-test.

### Sampling criterion and clinical culture for pathogen identification and antibiotic susceptibility testing

All urine samples collected in the study were initially screened for bacteriuria using a dipstick and urine flow cytometry on the UF-1000i system, as described by Fritzenwanker et al^48,^ Only those samples that tested positive for leukocytes and had appropriate cell counts from flow cytometry were included in the analysis. The university hospital’s microbiological laboratory performed routine microbiological analysis of the collected urine samples. In brief, 10 **µl** of urine was inoculated onto blood agar plates (CNA Blood, MacConkey, and CPSE) and incubated for 18-24 hours at 37°C in aerobic conditions. The identification of bacterial species and antibiotic susceptibility testing (AST) were performed separately using the MADI-TOF analysis (bioMerieux, Ntirtingen, Germany) and VITEK-2 systems (bioMerieux, Ntirtingen, Germany). The antimicrobial susceptibility test cards of the VITEK 2 AST-N432 and AST P611/654 Test Kits (bioMerieux, SA) were used for Gram-negative and Gram-positive organisms, respectively. The test results were interpreted according to EUCAST guidelines.

#### Testing of the 11 methods on complicated UTI samples

Eleven methods, comprising eight HL_SAN/M_SAN combinations and three commercial kits (Figure *1*), were evaluated on a combination of 77 patient samples collected from the Department of Urology, Pediatric Urology, and Andrology at Justus-Liebig University of Giessen in Germany. The clinical evaluation of the developed methods was carried out in two phases.

In the pilot phase, eight SAN methods (HL+NB, H+BT, M+NB, M+BT) and three commercial methods (MC5, HZ, and NB) were tested on clinical urine samples at the Institute of Medical Microbiology, Medical Microbiome - Metagenome Unit (M3U), Justus Liebig University Giessen, Giessen, Germany. All collected samples were one day old and tested positive for leukocytes and/or nitrate presence. Approximately 3-8 mL of each sample was collected and processed using the abovementioned methods without prior information on pathogen type or AST results. The samples were from a mix of male and female patients of various age groups and included some urosepsis samples. The extractions were performed in two rounds, with 15 samples processed and sequenced in round 1, followed by 12 samples in round 2.

After round 2, M+NB and H+NB protocols were modified to include either bead beating (BB) or enzymatic (LZ) lysis steps to enhance extraction efficiency for Gram-positive pathogens. In the enzymatic version (LZ) of the methodology, the samples post-depletion were treated with an enzymatic lysis buffer (20 mg lysozyme, 20 mM Tris.Cl, 2 mM sodium EDTA, and 1.2% Triton-X-100) and incubated at 37°C for 30 minutes before proceeding with the rest of the DNA extraction protocol from NB. In the bead-beating (BB) version, the samples post-depletion were first treated with 20 µL of proteinase K and incubated at 55°C for 10 minutes. The sample was transferred to Lysis Matrix E tubes and beaten at maximum intensity on a Vortex-Genie® 2 for 5 minutes. The lysis tubes were centrifuged at ≥ 10,000g for 60 seconds, and the supernatant was transferred to a fresh 2 mL Eppendorf tube before continuing with the remaining NB extraction steps. Subsequently, the best-performing method was selected based on a combination of host depletion, cost, and turnaround time. The optimized method was subsequently tested on 50 samples to evaluate its performance in comparison to the results of the clinical routine.

#### Vivalytic/PCR-based confirmation of additional pathogens from mNGS that were missed by the routine testing method

In addition to the routine culture analysis, some samples with additional bacteria detected metagenomically were reanalyzed using either Vivalytic or PCR based tests. Among samples 41,59, and 70, which had *M morganii, P stuartii*, and *P aeruginosa* detected metagenomically, retesting was conducted through Vivalytic, as described by Hartmann et al. For samples 53, 54, and 62, where *A. urinae* was identified, and samples 56, 57, and 62, where *A. schaalii* was found, and pathogen-specific primers were employed to confirm their presence.

#### Nanopore sequencing of the clinical samples

Post-extraction, all the extracted samples were purified and concentrated using the Agencourt AMPure XP system (Beckman Coulter, USA). Library preparation for MinION sequencing was conducted with the Rapid Barcoding Sequencing kit SQK-RBKl 14.24 (Oxford Nanopore, UK) and sequenced using MinION flow cells (Rl0.4.1 FLO-MIN 106D, Oxford Nanopore). Samples from both rounds were multiplexed and loaded separately onto two MinION flow cells.

#### Bioinformatics analysis of the sequencing data

The sequencing data were basecalled in real time using MinKNOW software (version 6.0.11) in fast basecalling mode with Dorado (version 7.4.13). Unclassified reads were recovered using MysteryMaster (Abdolrahman Khezri, SB, JA, ABB, Crystal Chapagain, Manfred Grabherr, RA) with default parameters. Reads were re basecalled with Dorado (version 0.6.4), employing HAC and SUP models to compare the results. Pathogen reads were identified using BLASTn against a custom reference genome panel of 63 common uropathogens, with low complexity regions masked using BBmask (BBMap version 38.84). BLASTn subsequently identified additional bacterial reads against the NCBI Prokaryotic Reference Genomes collection (RefProk). BLASTn was run with the following parameters: -word size 28 -max target seqs 150 - evalue 0.000001. The following cutoff values were used for bacterial identification: minimum percent identity: 80%, minimum read coverage in alignment: 65%, minimum read length: 200 nt. Reads that did not align with the RefProk database were discarded to remove human reads. If pathogen genome coverage exceeded 2%, reference-based assemblies were created using minimap2^54^ (version 2.22) with the ‘map-ont’ option. Samtools^55^ (version 1.13) was used to filter the alignments (options -b-F 4) and generate a consensus sequence. ARGs were identified using abricate^56^ (version 1.0.1) with the NCBI^57^ and CARD^58^ databases.

#### Statistical analysis

The area under the receiver operating characteristic curve (AUROC) was calculated using routine culture as the reference standard for total DNA yield and the number of bacterial cells per ml, as identified by flow cytometry. The optimal cutoff values were computed by maximizing Youden’s index^59^. Positive Predictive Value (PPV) and Negative Predictive Value (NPV) were calculated after classifying the data according to the optimal cutoff. The 95% confidence intervals for AUROC and PPV /NPV were determined through bootstrapping using 10,000 steps. The statistics described were calculated using the sklearn^60^ (vl .3.2) Python package.

## Data availability

The datasets presented in the study can be found through the accession number PRJEB83412 m the European Nucleotide Arc ive repository online usmg the URL: https://www.ebi.ac.uk/ena/browser/view/PRJEB83412

## Author contributions

RA, FW, TH, CI, and TBJ planned the study. ABB, SB, JA, and RA designed the experiments. ABB, JA, and FA performed the experimental work in discussions with RA. ABB, SB, and RA wrote the manuscript. All authors contributed to the article and approved the submitted version.

## Ethics information

The study’s Ethical approval was obtained from the Ethics committee of the Justus Liebig University Giessen, Faculty of Medicine (AZ 158/20).

