## Supplementary Figure 1 for "Accurate and rapid turnaround of four hours for diagnosis of complicated UTIs using metagenomics"

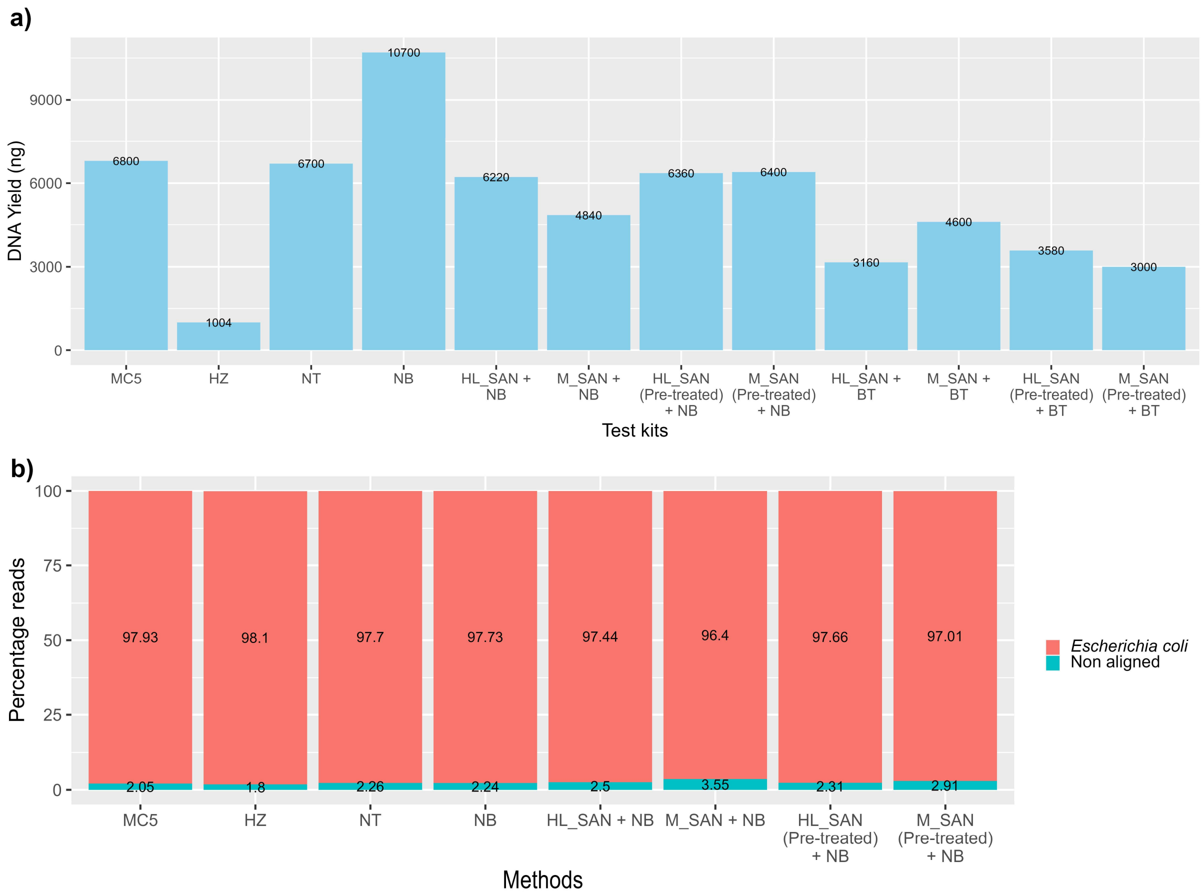


**Supplementary Figure 1:** a.) Bar graph denoting the DNA yield (ng) obtained from different DNA extraction methods tested on urine samples spiked with clinical *Escherichia coli* isolate. Sample treatment with EDTA (pre-treatment) had a minor effect on the DNA yield. b) The relative (read level) pathogen abundance from sequencing data for different DNA extraction methods. Reads were mapped against the NCBI prokaryotic reference genomes database. Reads that did not align to prokaryotic genomes were indicated as 'Nonaligned'. The names of the methods used are the same as in Supplementary Table 1. NT stands for Naxtra Total nucleic acid kit.
