## Supplementary Figure 2 for "Accurate and rapid turnaround of four hours for diagnosis of complicated UTIs using metagenomics"

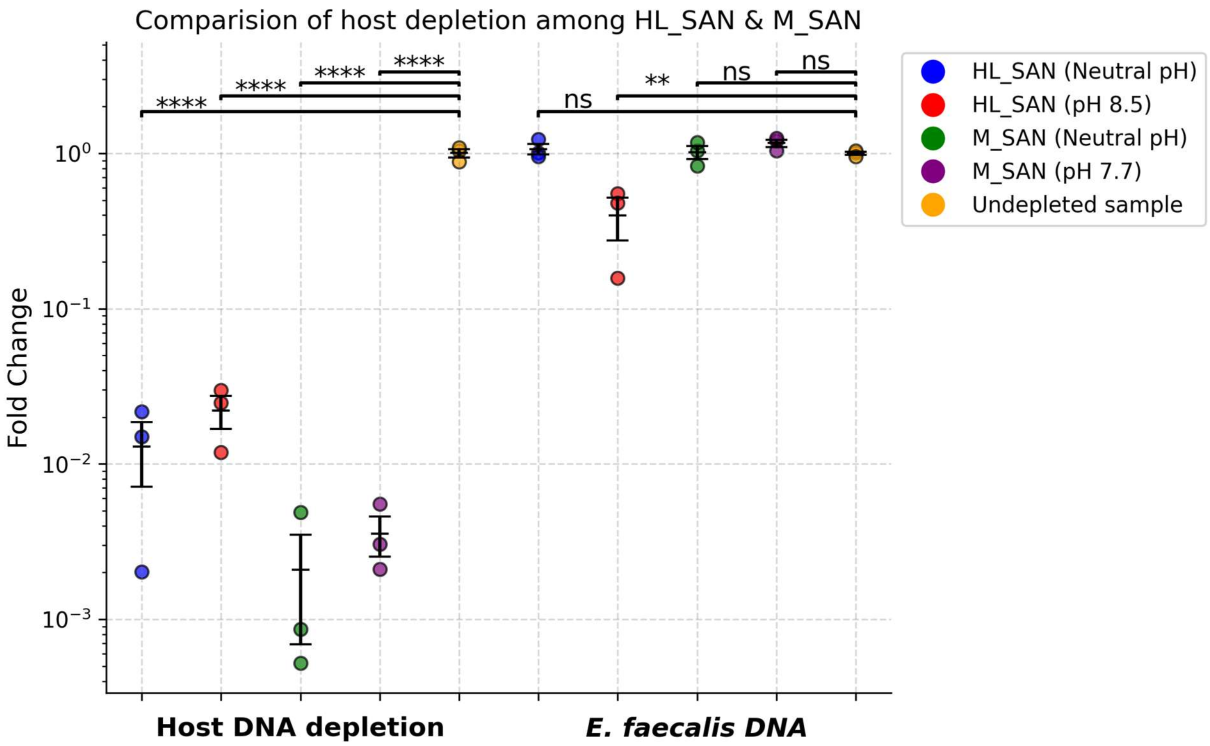


**Supplementary Figure 2:** Relative quantification of the host and the *Enterococcus faecalis* DNA in urine samples spiked with WBC and *E. faecalis* and subjected to depletion by HL_SAN and M_SAN at two different pH conditions to determine their respective depletion abilities. The values are normalized by determining the fold change calculated by the difference with the undepleted control. All the test values are mean ± SD, with n=3 biological replicates for each condition. ns > 0.05, *p ≤ 0.05, **p ≤ 0.01. ***p ≤ 0.001, ****p ≤ 0.0001.
