## Supplementary Figure 3 for "Accurate and rapid turnaround of four hours for diagnosis of complicated UTIs using metagenomics"

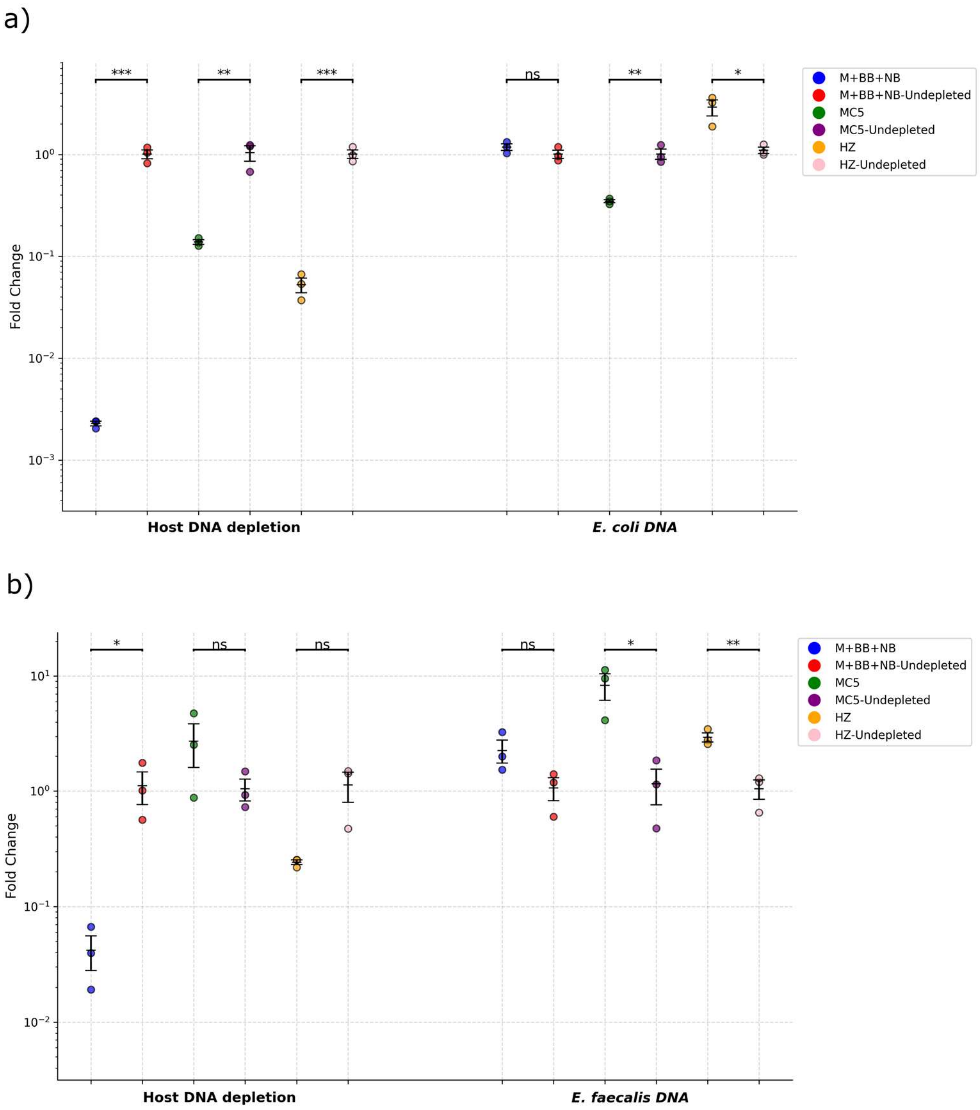


**Supplementary Figure 3:** Relative quantification of the host and the spiked bacterial DNA in urine samples spiked with WBC and subjected to depletion by different extraction methodologies to determine their respective depletion abilities. Two separate sample sets were spiked with *E. coli* (a) and *E. faecalis* (b) at clinically relevant concentrations of $10^5$ CFU/mL. The values are normalized by determining the fold change calculated by the difference with the undepleted control. All the test values are mean ± SD, with n=3 biological replicates for each condition. ns > 0.05, *p ≤ 0.05, **p ≤ 0.01. ***p ≤ 0.001, ****p ≤ 0.0001. Abbreviations: MC5: Molysis Complete 5, HZ: Host Zero Microbial DNA extraction kit, M+BB+NB: optimized in-house method.
