## Supplementary Figure 4 for "Accurate and rapid turnaround of four hours for diagnosis of complicated UTIs using metagenomics"

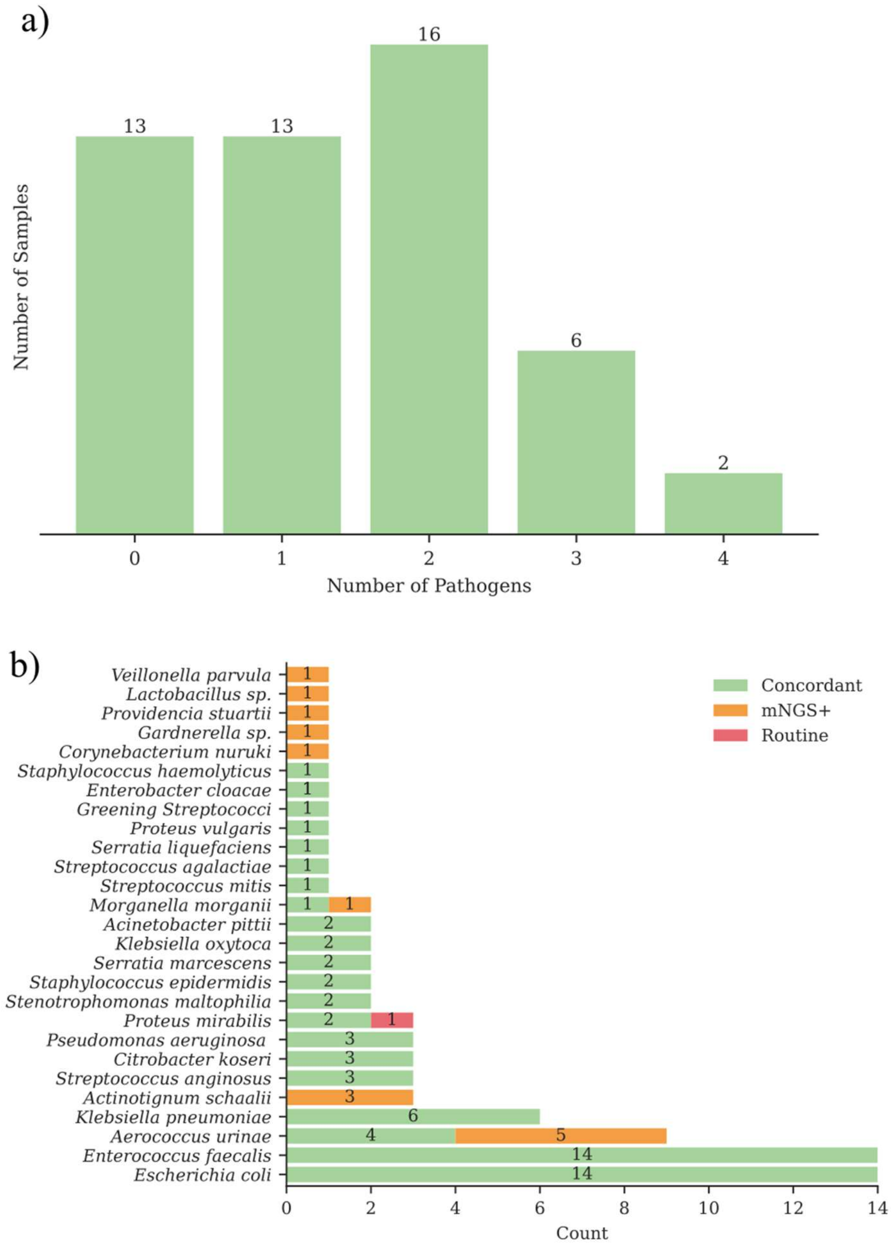


**Supplementary Figure 4:** Summary of the pathogen identification results for the in-house optimized method. (a) The histogram shows the number of samples with a certain number of pathogens, as identified through routine culturing. A value of 0 indicates culture-negative samples. Samples with one pathogen (13) were classified as mono-microbial, while those with more than one pathogen (24) were considered polymicrobial. (b) illustrates the frequency of identification for each species. Green bars represent samples where the species was found through both routine culturing and mNGS, orange bars denote samples identified solely through mNGS, and red bars indicate samples where the species was identified exclusively through routine culturing.
