## Supplementary Figure 5 for "Accurate and rapid turnaround of four hours for diagnosis of complicated UTIs using metagenomics"

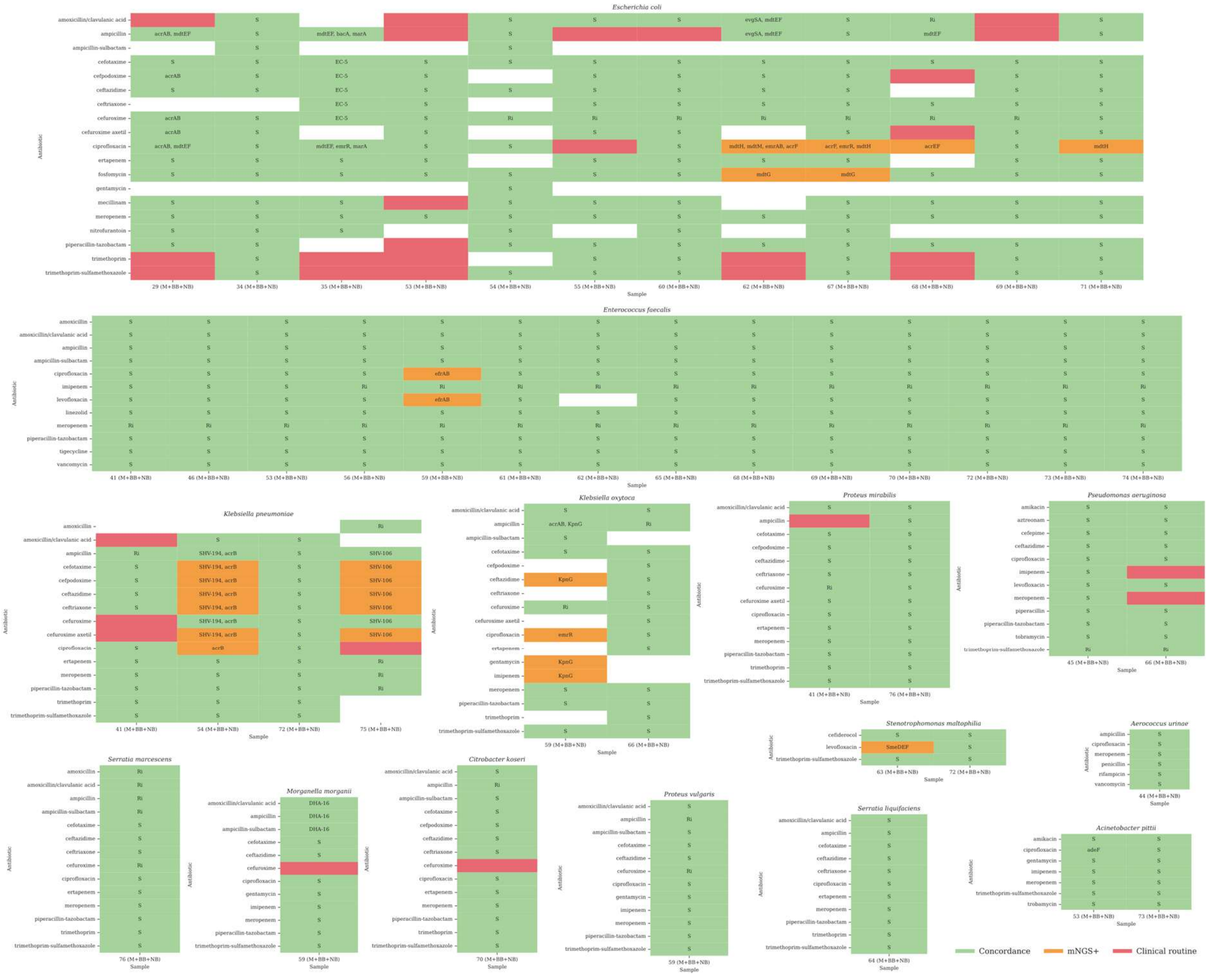


**Supplementary Figure 5:** Heatmaps showing the concordance between routine antimicrobial susceptibility testing (AST) results and metagenomic (mNGS) resistance predictions derived from antimicrobial resistance gene (ARG) data for samples processed using the optimized in-house method. The data are grouped by pathogen and displayed per antibiotic for each UTI sample. Green cells indicate concordance between observed phenotypes and detected ARGs (true positives or negatives). The cells are annotated with 'S' for susceptibility, 'Ri' for inferred or intrinsic resistance, and the ARG name when resistance mechanisms are detected. Red cells indicate instances where resistance was identified in routine AST, but no corresponding resistance mechanism was detected through mNGS (false negative). Orange cells represent detected ARGs without corresponding phenotypic resistance to the relevant antibiotic, and these cells are annotated with the ARG names. AMR predictions were made exclusively for antibiotics present in both the phenotypic and genotypic datasets.
