## Supplementary Figure 6 for "Accurate and rapid turnaround of four hours for diagnosis of complicated UTIs using metagenomics"

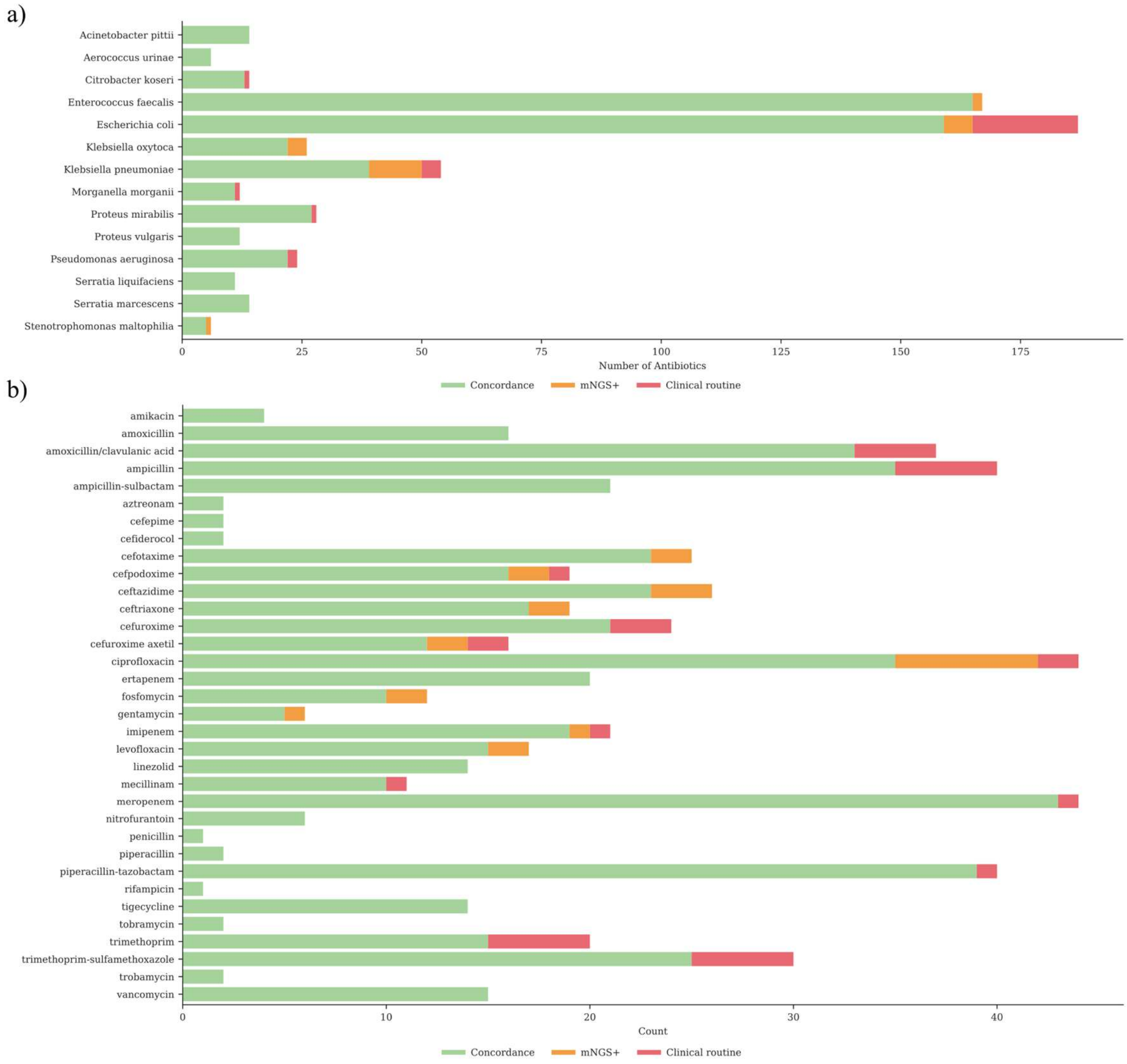


**Supplementary Figure 6:** Overview of the concordance between routine antimicrobial susceptibility testing (AST) results and metagenomic (mNGS) resistance predictions derived from antimicrobial resistance gene (ARG) data for samples processed using the optimized in-house method. Green bars indicate concordance between observed phenotypes and detected ARGs (true positives or negatives). Red bars highlight instances where resistance was identified in routine AST, but no corresponding resistance mechanism was detected through mNGS (false negatives).  Orange bars signify detected ARGs without phenotypic resistance to the corresponding antibiotic. AMR predictions were made exclusively for antibiotics present in both the phenotypic and genotypic datasets.
