## Supplementary Figure 7 for "Accurate and rapid turnaround of four hours for diagnosis of complicated UTIs using metagenomics"

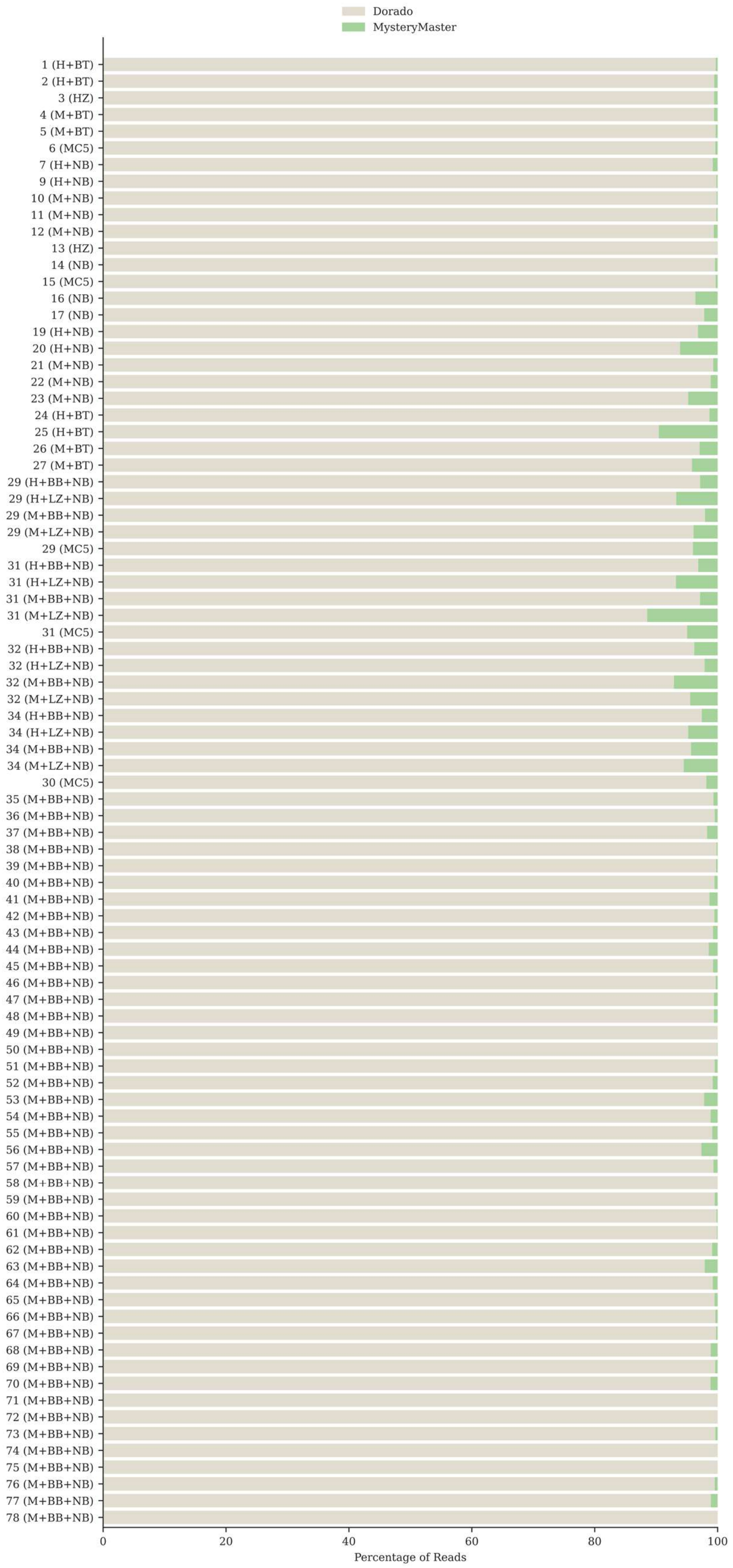


**Supplementary Figure 7:** Bar graphs illustrating the percentage of sequencing reads classified by Dorado and those unclassified, which MysteryMaster recovered. Green indicates the reads recovered by MysteryMaster.
