## Supplementary Figure 8 for "Accurate and rapid turnaround of four hours for diagnosis of complicated UTIs using metagenomics"

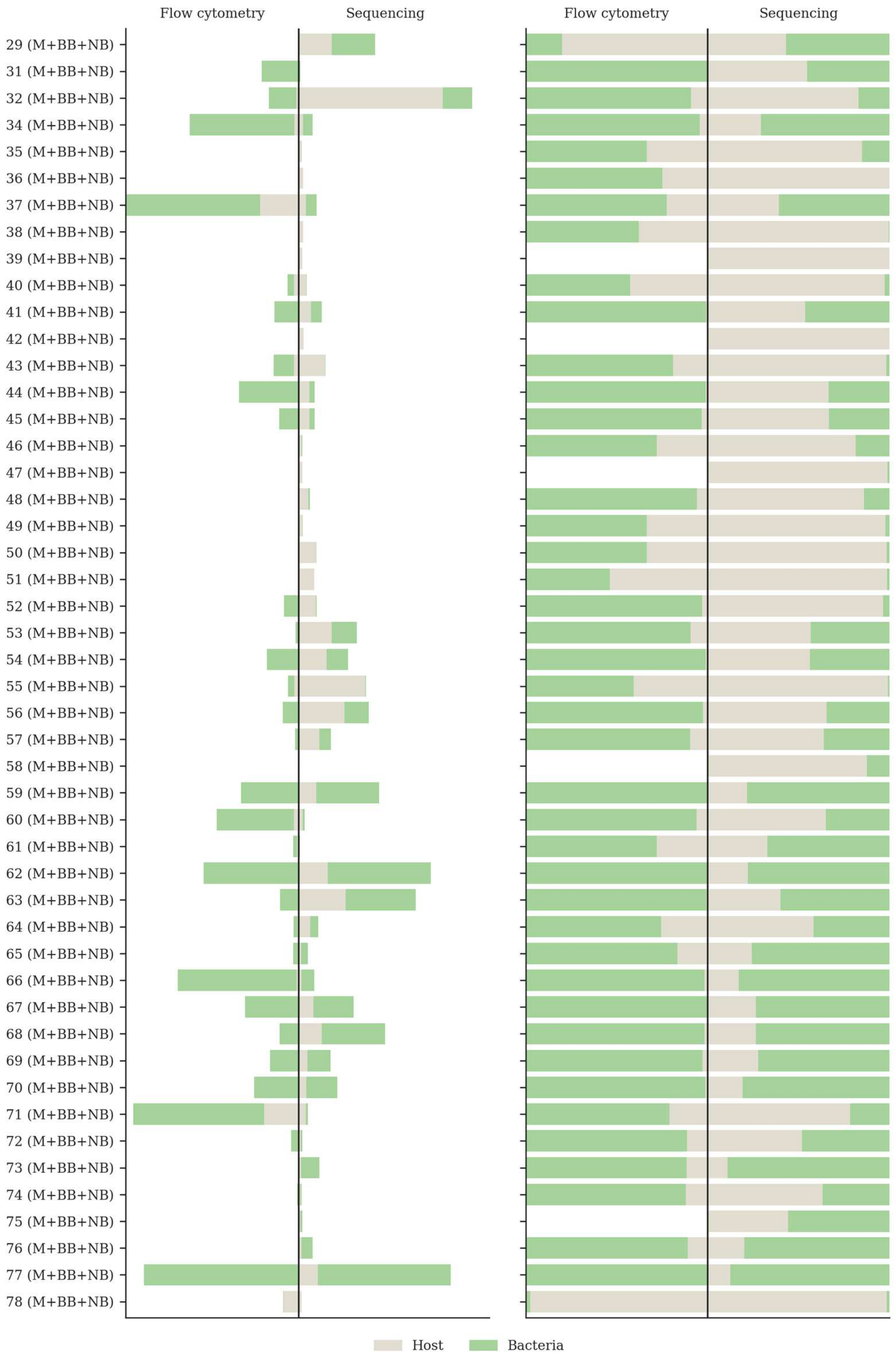


**Supplementary Figure 8:** The tornado charts illustrate the relationship between host (in grey) and bacterial cells (in light green) based on flow cytometry data, compared to host-to-bacterial reads obtained through metagenomic next-generation sequencing (mNGS). The left panel displays the normalized data, while the right panel presents the absolute data. Together, these plots enhance our understanding of the effectiveness of various methodologies across a range of clinical samples for the in-house optimized method.
