## Supplementary Figure 9 for "Accurate and rapid turnaround of four hours for diagnosis of complicated UTIs using metagenomics"

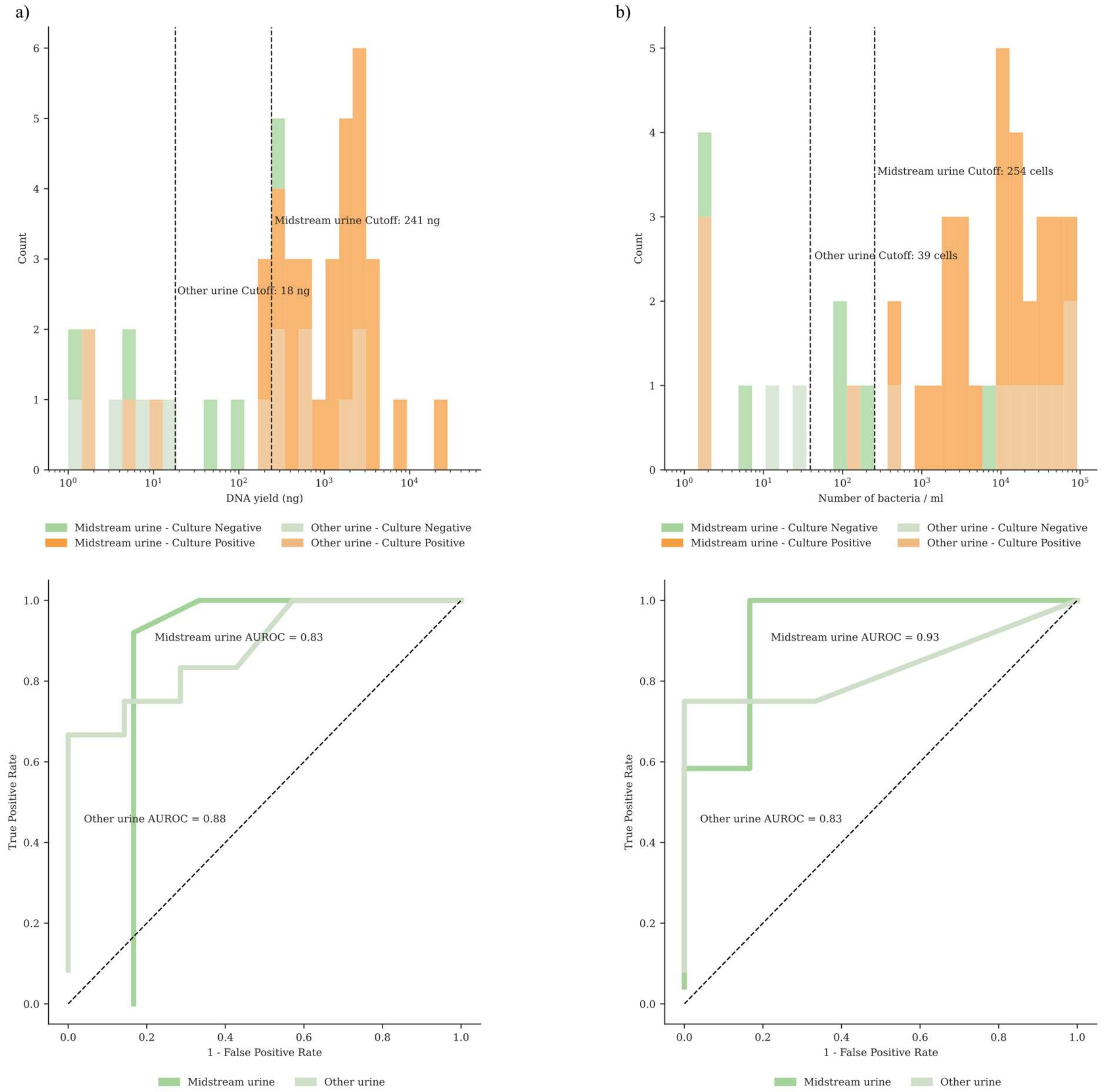


**Supplementary Figure 9:** Analysis of DNA yield and bacterial cell receiver operating characteristic (ROC) curve, divided by midstream urine and other urine. The analysis was performed on samples extracted using the in-house optimized method (n=50). Subfigure (a) presents the results for DNA yield, while (b) displays the bacterial cell count per mL identified through flow cytometry. The histogram (top) illustrates the distribution of each variable for culture-positive and culture-negative samples, emphasizing the optimal cutoff for distinguishing between these samples. Colors indicate whether the samples are from midstream urine or other urine. The ROC curve (bottom) is annotated with the area under the ROC curve (AUROC) for each variable.
