## Supplementary Figure 10 for "Accurate and rapid turnaround of four hours for diagnosis of complicated UTIs using metagenomics"

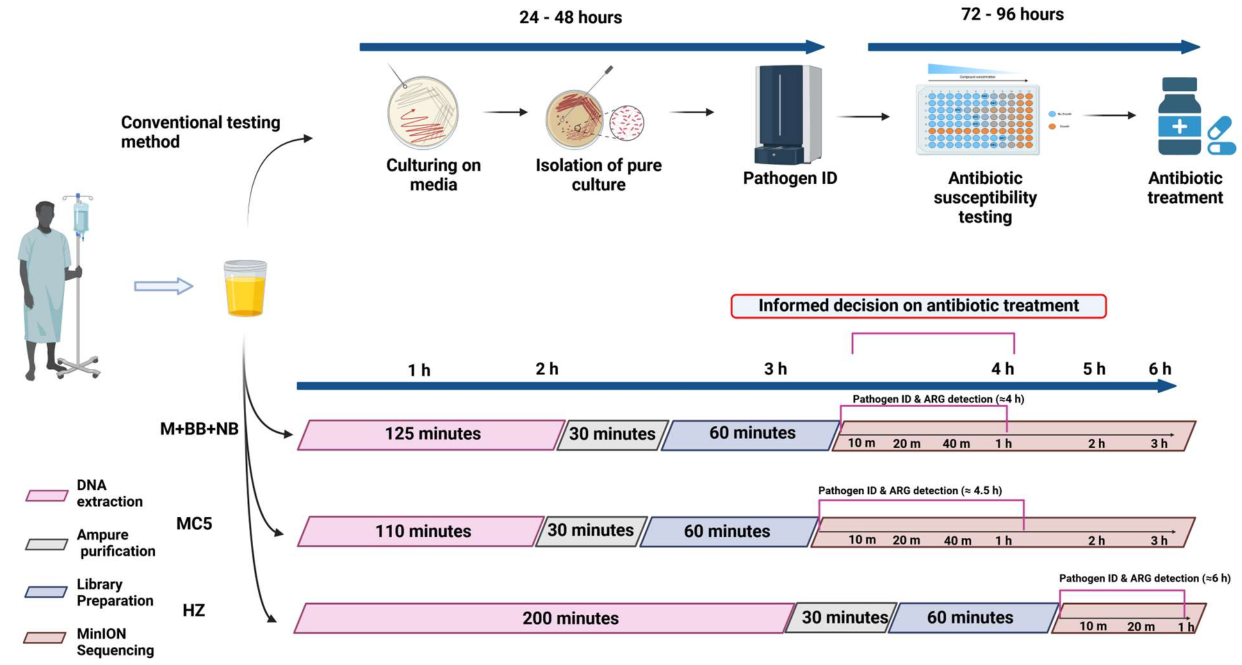


**Supplementary Figure 10:** Comparison of the total turnaround time between the in-house optimized method (M+BB+NB) and the commercial methods (MC5 and HZ) for the clinical samples. The timeline for the method is divided and color-coded according to DNA extraction, purification, library preparation, and sequencing. Overall, the optimized method achieved a turnaround time of approximately 4 hours for determining the antibiotic treatment. In contrast, MC5 and HZ had turnaround times of 4.5 hours and 6 hours, respectively.
