## Supplementary Table 1 for "Accurate and rapid turnaround of four hours for diagnosis of complicated UTIs using metagenomics"

**Supplementary Table 1:** An overview of all the different methods used in the study. The method numbering is the same as shown in Figure 1.

| Method number | Method combination | Abbreviation | Method category | Endonucleases used | Cell lysis combination | DNA extraction method |
|---|---|---|---|---|---|---|
| 1 | Molysis Complete 5 | MC5 | Commercial | - | According to the standard protocol | According to the standard protocol |
| 2 | Host Zero Microbial DNA extraction kit | HZ | Commercial | - | According to the standard protocol | According to the standard protocol |
| 3 | Naxtra Blood Total nucleic acid kit | NB | Commercial | - | According to the standard protocol | According to the standard protocol |
| 4 | HL_SAN + Blood and Tissue | H + BT | In-house | HL_SAN | Enzymatic lysis | Column based |
| 5 | M_SAN + Blood and Tissue | M + BT | In-house | M_SAN | Enzymatic lysis | Column based |
| 6 | HL_SAN + Naxtra Blood | H + NB | In-house | HL_SAN | Chemical lysis | Magnetic beads |
| 7 | M_SAN + Blood and Tissue | M + BT | In-house | M_SAN | Chemical lysis | Column based |
| 8 | HL_SAN + Enzymatic lysis + Naxtra Blood | H + LZ + NB | In-house | HL_SAN | Enzymatic lysis + Chemical lysis | Magnetic beads |
| 9 | HL_SAN + Bead beating + Naxtra Blood | H + BB + NB | In-house | HL_SAN | Mechanical lysis + Chemical lysis | Magnetic beads |
| 10 | M_SAN + Enzymatic lysis + Naxtra Blood | M + LZ + NB | In-house | M_SAN | Enzymatic lysis + Chemical lysis | Magnetic beads |
| 11 | M_SAN + Bead beating + Naxtra Blood | M + BB + NB | Optimized in-house | M_SAN | Mechanical lysis + Chemical lysis | Magnetic beads |
